# Gut microbiome alterations predictive of cancer risk

**DOI:** 10.64898/2026.09.09.26362628

**Authors:** Giacomo Vitali, Johannes R. Björk, Oleg Kambur, Marine Fidelle, Einar Birkeland, Lindsay Goulet, Emmanuelle Le Chatelier, Yani Ren, Robert Benamouzig, Patrick Veiga, Guido Kroemer, Trine Ballestad Rounge, Barbara Pardini, Teemu Niiranen, Mārcis Leja, Juozas Kupčinskas, Bertrand Routy, Arielle Elkrief, Joana Ribeiro, Lisa Derosa, Nikos Paragios, Jan Baumbach, Leo Lahti, Le French Gut Consortium, Mathieu Almeida, Rinse K. Weersma, Laurence Zitvogel, Stanislav Dusko Ehrlich

## Abstract

Cancer is the second leading cause of death worldwide. A large proportion of cancer deaths is associated with modifiable risk factors, such as tobacco use and dietary factors. This highlights the critical role of prevention to reduce cancer deaths, which relies on both identifying individuals at risk and mitigating that risk. Here, we focus on risk stratification by leveraging gut microbiome alterations. Utilizing two cancer discovery cohorts and four general population cohorts across different European countries - encompassing 16,255 individuals, including 3,341 cancer cases - we first establish the existence of common gut microbiome alterations across multiple distinct malignancies (lung, kidney, and breast). We demonstrate that these alterations are conserved in populations from France, Finland, the Netherlands, and Latvia. Crucially, we show that individuals who develop cancer after microbiome sampling (n = 1,642) harbor microbiome alterations similar to those found in clinically diagnosed cancer cases, albeit to a lesser degree. This indicates that the gut microbiome evolves towards a cancer-associated state well before clinical diagnosis. Finally, we demonstrate that the extent of microbiome alterations can effectively stratify both the patients with diagnosed cancer and individuals at elevated cancer risk.

## Introduction

In Europe, there are an estimated 4.1 million new cancer cases and about 2 million cancer deaths each year. Cancers of the breast, colon or rectum, lung, and prostate are the most common, representing half of the overall cancer burden (Ferlay et al., 2024). The 2026 US cancer statistics (Siegel et al., 2026) highlights the progress of modern oncology, i.e a drop in the overall cancer mortality rate over the last 32 years with a current 5-year relative survival rate for all cancers of 70%. These combined metrics validate the investment in tobacco control, human papilloma virus vaccines, early cancer detection strategies and an expanding oncological armamentarium. However, beyond this apparent success lies the need for further preventing cancers and improving survival, and addressing a number of persistent limitations, including national averages that mask disparities related to stage, cancer histology, and population specificities (ethnicity, socioeconomical status, and remote locations). Hence, modernizing surveillance and screening approaches, expanding coverage infrastructure, and scaling proven prevention equitably remain critical factors to pursue this quest to reducing cancer incidence and mortality rates worldwide (Zhang & Zhang, 2026). Effective screening paradigms exist only for a small subset of cancers, are focused on single cancer types, and have variable adoption and compliance.

Susceptibility to cancer is multifactorial. In addition to genetic risk factors, as indicated by risk stratification using polygenic risk scores (Yang et al., 2023), several lifestyle, anthropometric, hormonal, reproductive and imaging risk factors are known to be associated with risk for developing various cancers.

Behavioral, lifestyle and environmental risk factors associated with cancer susceptibility may translate into measurable biological features of potentially high clinical significance. The transition from health to disease may involve local organ-specific and systemic inflammation that is governed and maintained by the intertwined relationship between somatic mosaicism (Coorens et al., 2025), clonal hematopoiesis of indeterminate potential (Jaiswal et al., 2014), dysmetabolism and maladaptive immunity (Pennycuick et al., 2020), mostly controlled by age-related failure of the autophagy machinery (López-Otín et al., 2023; López-Otín & Kroemer, 2021; Nighot et al., 2025).

Over the last fifteen years, the integrity of the intestinal barrier and its natural commensalism, the gut microbiome, have emerged as critical determinants in the transition of health to chronic inflammatory disorders (Ghosh et al., 2022; Goel et al., 2025; Grajeda-Iglesias et al., 2021; Porcari et al., 2025; Stanley et al., 2016; Zitvogel et al., 2025). Indeed, intestinal dysbiosis, defined as an imbalance of the composition or function of the gut microbiome, is causatively linked to lymphocyte functional exhaustion associated with key metabolic perturbations (Zitvogel et al., 2024), culminating in resistance to cancer immunotherapy, in particular immune checkpoint inhibitors (ICI) and CAR-T cells (Routy, Gopalakrishnan, et al., 2018; Routy, Le Chatelier, et al., 2018; Thomas et al., 2023). More than half of patients with advanced cancer suffer from gut dysbiosis (Derosa et al., 2024), caused by cancer-related stress ileopathy (Yonekura et al., 2022), co-morbidities and comedications including antibiotics (Routy, Le Chatelier, et al., 2018; Thomas et al., 2023). Gut oncomicrobiome signatures have been described in network programs such as ONCOBIOME (Zitvogel et al., 2025) across various malignancies and are currently used as prognosis biomarkers in advanced cancers amenable to immunotherapy-based regimens (Almonte et al., 2026; Gopalakrishnan et al., 2018; Thomas et al., 2023). However, to which extent microbiota taxonomic deviations could precede cancer incidence and represent potential biomarkers of cancer risk has never been established.

An innovative approach to microbiome analysis, focusing on coordinated shifts of microbial species, has recently been applied to the study of Parkinson’s disease (Menozzi et al., 2026). It is based mainly on identifying coherent microbiome associations by comparisons of effect sizes across datasets, using the robust and non-parametric measure Cliff’s delta (Cliff, 1993; Meissel & Yao, 2024). This measure is increasingly reported in microbiome studies, alongside statistical significance of differential abundance of a microbial species between the groups of individuals. It describes the dominance of abundance in one group relative to another and is much less sensitive to outliers than the more widely used fold change parameter. Furthermore, for low values, its distribution tends to be normal and can thus be analyzed by parametric statistical tools, which are more powerful than the non-parametric ones usually deployed in microbiome analysis. However, until recently, Cliff’s delta has not been employed to assess the coherence of microbiome alterations between different cohorts of individuals, with a single exception of cardio-vascular ischemic disease (Fromentin et al., 2022). This analytical framework has recently been greatly extended to evaluate coordinated gut microbiome species shifts in neurodegeneration (Menozzi et al., 2026), demonstrating the feasibility of disease-risk stratification.

Following this lead, we analyzed the gut microbiome of individuals with cancers diagnosed *prior to/at the time of* or *after* microbiome sampling. The former group allowed assessment of microbiome alterations related to cancer presence and common to different cancers. The latter group, considered at risk of cancer because they received a cancer diagnosis after sampling, allowed assessment of whether similar microbiome alterations can be detected before cancer diagnosis, and thus be indicative of progression towards cancer. Here, we report microbiome alterations common to different cancer types, which are present in individuals with no diagnosed cancer. We show that stratification of these individuals into groups at higher and lower cancer risk can be achieved by measuring the extent of coherent gut microbiome alterations.

## Results

### Study populations and analytical strategy

We used two cohorts of French cancer patients, Canto (n=83) and Oncobiotics (n=637), focusing on early breast cancer for the former and mostly advanced and metastatic lung (n=585) cancer for the latter, but including kidney (n=38) and bladder cancers (n=14) as well (**Supplementary Table 1a, b)**. We used four European population cohorts. (i) LifeLines from the Netherlands, including n=248 cancer cases diagnosed before microbiome sampling (hereafter termed “prevalent cancers”), n=245 cancer cases diagnosed after sampling (hereafter termed “incident cancers”) and n=701 cancer-free controls; (ii) FINRISK from Finland including prevalent and incident cancer cases (n=268 and n=1105, respectively) and n=2525 cancer-free controls; (iii) French Gut from France including prevalent and incident cancers (n=463 and n=86, respectively) and n=9437 cancer-free controls and (iv) GISTAR from Latvia including incident cancers (n=206) and cancer-free controls (n=251). Relevant cohort details are reported in Table 1 and **Supplementary Table 1c-f**.

**Table 1.** Cohort characteristics.

| Characteristic | Immunolife, France<br>N = 637<sup>1</sup> | Canto breast,<br>France<br>N = 83<sup>1</sup> | French Gut, France<br>N = 9,986<sup>1</sup> | Lifelines,<br>Netherlands<br>N = 1,194<sup>1</sup> | Finnrisk, Finland<br>N = 7,231<sup>1</sup> | Gistar, Latvia<br>N = 455<sup>1</sup> |
| --- | --- | --- | --- | --- | --- | --- |
| Age, years | 65.0 (58.0, 71.0) | 52.0 (47.0, 62.0) | 47.0 (34.0, 59.0) | 51.0 (43.0, 60.8) | 49.5 (41.1, 57.4) | 58.0 (52.0, 63.0) |
| Missing | 1 | 0 | 33 | 0 | 0 | 0 |
| Sex |  |  |  |  |  |  |
| F | 222 (34.9%) | 83 (100.0%) | 6,903 (69.1%) | 604 (50.6%) | 3,979 (55.0%) | 237 (52.1%) |
| M | 414 (65.1%) | 0 (0.0%) | 3,083 (30.9%) | 590 (49.4%) | 3,252 (45.0%) | 218 (47.9%) |
| Missing | 1 | 0 | 0 | 0 | 0 | 0 |
| BMI, kg/m<sup>2</sup> | 23.9 (21.1, 26.9) | 25.8 (21.9, 29.0) | 23.2 (20.9, 26.3) | 25.1 (23.0, 27.2) | 26.3 (23.3, 29.6) | 28.1 (24.6, 32.2) |
| Missing | 6 | 46 | 0 | 0 | 2 | 2 |
| Cancer |  |  |  |  |  |  |
| control | 0 (0.0%) | 0 (0.0%) | 9,437 (94.5%) | 701 (58.7%) | 5,858 (81.0%) | 251 (55.2%) |
| incident_cancer | 0 (0.0%) | 0 (0.0%) | 86 (0.9%) | 245 (20.5%) | 1,105 (15.3%) | 204 (44.8%) |
| prevalent_cancer | 637 (100.0%) | 83 (100.0%) | 463 (4.6%) | 248 (20.8%) | 268 (3.7%) | 0 (0.0%) |

<sup>1</sup> Median (Q1, Q3); n (%)

The overall analytical strategy is summarized in Figure 1. Cancer discovery cohorts were used to explore and define common cancer-related gut microbiome alterations by comparing them with cancer-free French Gut controls matched by age, sex, and BMI. Population cohorts were then used to explore whether (i) similar alterations can be detected when comparing prevalent cancer cases with healthy controls from the same cohorts, and (ii) whether incident cancer cases diagnosed after microbiome sampling carry alterations similar to those of prevalent cancers. The level of microbiome alteration was assessed and used to distribute individuals into quintiles, allowing for the computation of relative cancer risk by comparing the extreme quintiles.

**Fig 1.**
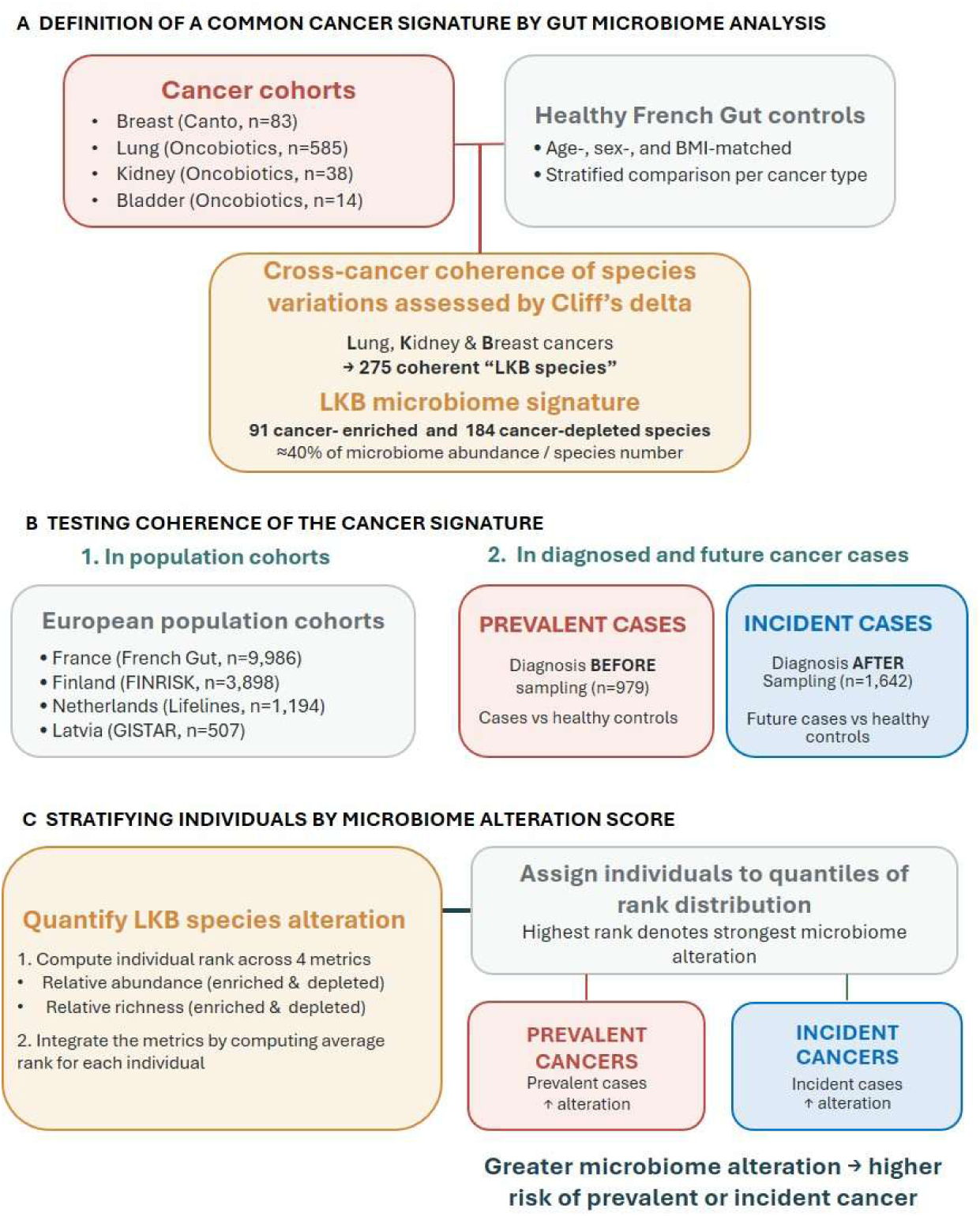
Analytical strategy: from common cancer-associated microbiome changes to cancer-risk stratification. (A) Identification of a cross-cancer microbiome signature. Metagenomic datasets from discovery cancer cohorts (Breast, Lung, Kidney, and Bladder) are compared against age-, sex-, and BMI-matched healthy controls (French Gut) using Cliff’s delta. A shared cross-cancer signature is defined by the coherence of species variations across Lung, Kidney, and Breast (LKB) cancers, yielding a core set of 275 LKB species, characterized by patterns of enrichment and depletion in cancer relative to controls. (B) Large-scale assessment across European population cohorts. The coherence and robustness of the LKB microbiome signature are independently tested within four distinct population cohorts (France, Finland, Netherlands, and Latvia). The analysis evaluates both prevalent cases (individuals diagnosed before fecal sampling; n = 979) and incident cases (future cancer cases diagnosed after fecal sampling; n = 1,642), comparing each against their respective healthy controls. (C) Individual stratification by microbiome alteration score. Individual LKB species alteration is quantified by computing ranks across four metrics capturing relative abundance and richness for both enriched and depleted species. Individual ranks are integrated into an overall average rank to distribute individuals into quantiles. Higher average ranks reflect stronger microbiome alteration, indicating a greater risk of prevalent or incident cancer.

The population cohort controls were consistently non-smokers with a BMI below 30 and free of cancer and other chronic diseases. This design aimed to accentuate microbiome differences between controls and cancer cases, given the heterogeneity of the cases with respect to cancer type, likely severity, and time of diagnosis relative to microbiome sampling, likely by dampening inflammatory baseline of controls. While such an unconventional approach to case-control comparisons could potentially introduce microbiome differences driven by factors other than cancer, we addressed this concern by focusing strictly on cancer-related differences. These were identified by their consistency with the common cancer-related alterations revealed in the initial comparison between cancer cases and matched cancer-free controls.

### Gut microbiome alterations common to different cancers

To search for species altered in cancer relative to healthy individuals, we compared the microbiomes of Oncobiotics lung, kidney and Canto breast cancer patients with those of French Gut controls, matched by age, sex, and BMI, when available, and distinct for each cancer type. Cliff’s delta values were computed for species of the Meteor2 database (n=2,742), which were common to cancer cases and healthy controls in each comparison. Cliff deltas correlated positively for all species in the three comparisons (**Fig. 2A-C**; the number of species is indicated in the panels). Correlation coefficients increased substantially when analyses were restricted to species that differed in abundance between healthy and cancer patients at p < 0.05 by Wilcoxon test (**Fig. 2D-F**). Significantly more species were enriched or depleted coherently than non-coherently in the three comparisons (**Fig. 2 G-I;** *χ^2^ test* values varied between 10^-4^ and 10^-14^ as indicated in cognate panels). These observations indicate that microbiome alterations in different cancer-types compared to healthy individuals are similar.

**Fig 2.**
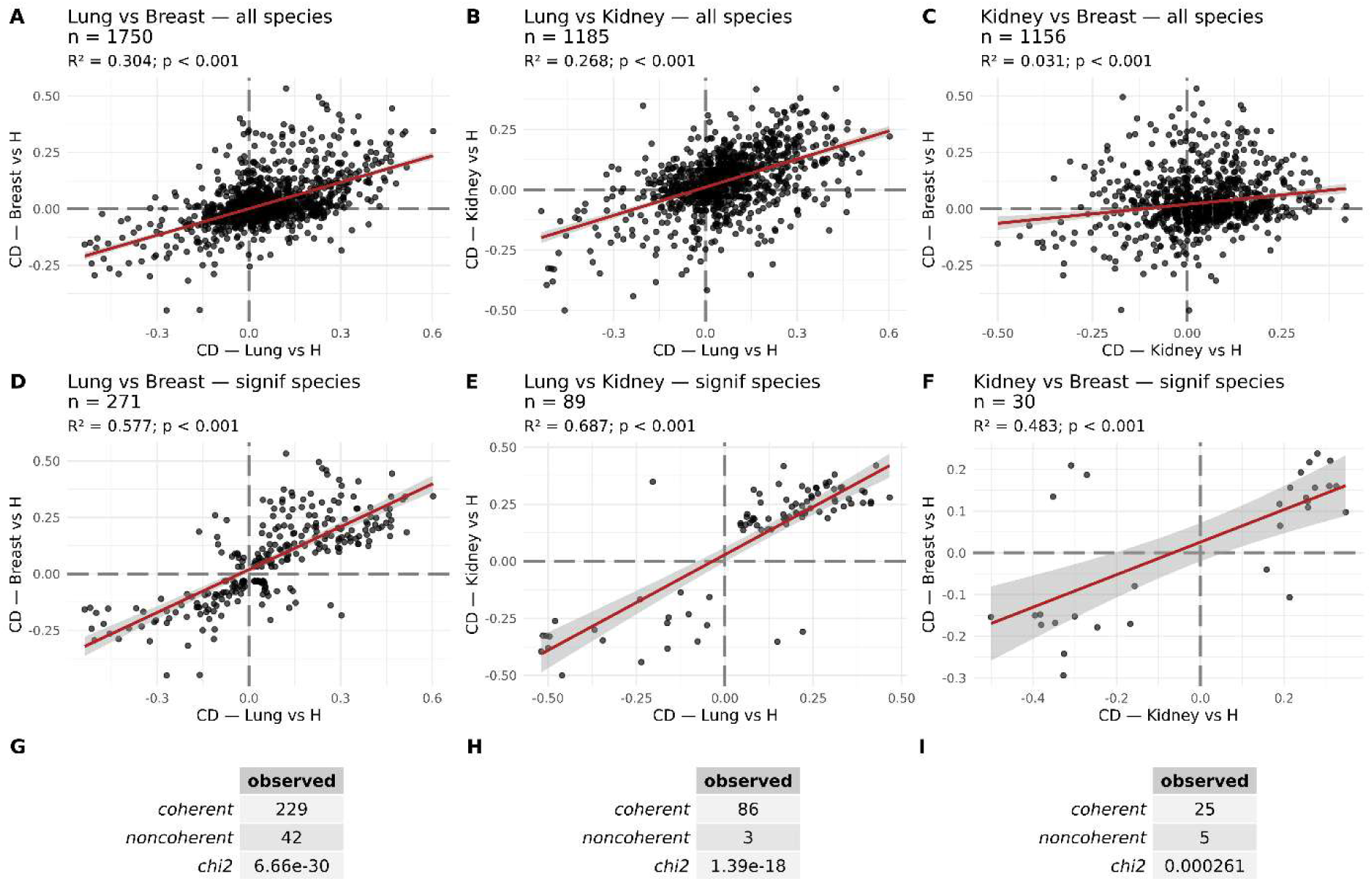
Coherence of gut microbiome alterations in lung, breast and kidney cancers. Cancer cases were matched with healthy individuals from the French Gut cohort by age, sex, and BMI for lung (n=585) and kidney (n=38) cancers and for age and sex for breast cancers (n=83, BMI data were not available) and cliff delta values were computed for 2742 species of the Meteor v2 catalogue. Cliff deltas were compared pairwise, for lung and breast cancers (A and D), lung and kidney cancers (B and E) and kidney and breast cancers (C and F), for all common species (A-C) or species with significantly different abundances (Wilcoxon p<0.05; D-F) in comparison between healthy controls and cancer cases; the number of species for each comparison is indicated in the corresponding panels. The number of species with significantly different abundances that varied coherently and non-coherently is indicated in panels G-I, together with the significance of the imbalance of coherent and non-coherent species, estimated by the *χ^2^* test. R^2^ denotes the squared Pearson correlation coefficient, n refers to number of species in each comparison, p denotes significance of correlation.

To further explore coherence of microbiome alterations, we focused on 498 species detected in at least 10% of lung cancer patients, our largest group, and differing significantly in abundance from healthy controls (Wilcoxon test p<0.05). A majority of these species (55%, n=275; **Supplementary Table 2**) was coherently enriched (n= 91) or depleted (n=184) also in kidney and breast cancers and showed a highly correlated Cliff’s delta (Pearson R^2^= 0.3 to 0.64) in the three comparisons (**Supplementary Fig 1**), although most (94.2%) did not reach a significance of Wilcoxon test p<0.05 in all three comparisons. This indicates that coherent microbiome alterations in cancer may involve many more species than detected when restricting the analysis to species that individually display statistically significant differences in abundance. To test this conclusion further, we examined coherence of Cliff’s delta variations of the 275 species in an unrelated cancer, stage III bladder neoplasia from the Oncobiotics cohort (n=14) matched by age, BMI, and sex to healthy French Gut controls. Of the 275 species coherently altered in lung, kidney and breast cancers, 179 (65%) were coherently altered in bladder cancer (*χ^2^ test* p=2.4E-17), indicating similar microbiome alterations in yet another cancer type, notwithstanding the small sample size.

To explore the potential confounding effects of antibiotic treatment (commonly undergone by patients with cancer) and smoking (frequent among lung cancer patients), we matched a sub-cohort of Oncobiotics lung cancer patients who had not received antibiotics (n = 440) with healthy controls based on BMI, age, sex, and smoking status (current, former, or never smoker). We then computed Cliff’s delta for all species present in this sub-cohort (n = 1,711). These included all LKB species, for which the correlation between the Cliff’s delta values computed using the entire lung cancer cohort (matched by BMI, age, and sex) and those from the antibiotic-free, smoking-matched sub-cohort was nearly perfect (Pearson R^2^ = 0.99, slope = 0.97; data not shown) with complete directional coherence (91 enriched and 184 depleted species). Based on these results, we conclude that antibiotics and smoking did not confound the identification of cancer-related species in our study.

Based on these observations, we propose that 275 species coherently altered in Oncobiotics lung (L) and kidney (K) cancers and Canto breast (B) cancers (hereafter designated as “LKB species”), might be used to assess microbiome alterations in different cancer types. They represent collectively almost 40% of the total abundance and number of the species within an individual’s microbiome, (**Supplementary Fig 2 A, D**) suggesting potential for extensive alterations of the microbiome across different cancer types. Expectedly, enriched and depleted species represented significantly higher and lower proportion of the microbiome in cancer cases than in healthy controls, respectively, both by relative abundance and relative number (**Supplementary Fig 2 B, C, E, F**). Notably, cancer-enriched species have lower overall abundance and number than cancer-depleted species, in particular in healthy individuals (5-fold and 3-fold lower, respectively, **Supplementary Fig 2, B, C**) and their increase in cancer is proportionally substantially higher (up to 2.2-fold by abundance and 2-fold by number) than the decrease of depleted species (at most 0.7-fold by abundance or number).

Interestingly, the microbial species enriched in cancer cases encompass many bacterial representatives included in the SIG1 features of the topological score TOPOSCORE (Derosa et al., 2024), associated with intestinal inflammation (*Ruminococcus gnavus* and *Mediterraneibacter torques* (Hall et al., 2017), autoimmune inflammation (*Eisenbergiella tayi,* (Yoon et al., 2025) or immunoresistance in immune checkpoint inhibition (such as *Eggerthella lenta, Bacteroides uniformis, Enterocloster spp, Parabacteroides distasonis, Flavonifractor plautii, Limosilactobacillus fermentum* (Birebent et al., 2025; Derosa et al., 2022, 2024), or antibiotic exposure and severe dysbiosis associated with poor prognosis, such as *Hungatella spp., Otoolea spp.,* and oral taxa (Derosa et al., 2020; Hakozaki et al., 2020). Conversely, microbial species depleted in cancer patients compared with healthy individuals belong to short-chain fatty acid (SCFA) producers and SIG2 features of the TOPOSCORE, such as *Dorea longicatena, Coprococcus catus, Akkermansia muciniphila, Roseburia yibonii, Ruminococcus callidus, Faecalibacterium spp, Eubacterium ventriosum* and other Eubacterium species*, Alistipes communis* and distinct *Lachnospiraceae, Oscillibacter and Oscillispiraceae spp.* (Birebent et al., 2025; Derosa et al., 2022, 2024). The collective coordinated shift of cancer-enriched and -depleted taxa constitutes a distinct signature across evaluated malignances, notwithstanding their association with systemic or barrier inflammation.

### Gut microbiome alterations common to cancers in different populations

We next explored the variation of LKB species in different population cohorts. To this end, we computed Cliff’s delta in the Lifelines Dutch Microbiome cohort, comparing healthy individuals (n=701) with prevalent pan-cancer cases (n=248). A total of 524 species were identified at a prevalence ≥10% by MetaPhlAn4 (Blanco-Míguez et al., 2023) in the cohort and 415 (79.1%) were matched by taxonomic assignments to species of Meteor2 database, including 159 LKB species. Of these, 128 (81%) were enriched or depleted in the same direction as in lung, kidney and breast cancers used to define the LKB species (**Supplementary Table 3**), a very strong bias towards coherence (*χ*^2^ test p=1.4E-14). Furthermore, Cliff’s deltas between LKB and LifeLines cancers were highly correlated (Pearson R2= 0.3 to 0.6, **Supplementary Fig 1, D-F**). Among the cancer-depleted species commonalities, we found *Dorea longicatena, Eubacterium ventriosum, Akkermansia muciniphila, Alistipes communis, Ruminococcus callidus, Roseburia yibonii,* among other prevalent SIG2 and/or immunogenic bacteria (**Supplementary Tables 2 and 3)**. Among the shared cancer-enriched species commonalities, we identified *Mediterraneibacter torques, Eggerthella lenta, Bacteroides uniformis, Parabacteroides distasonis,* and *Flavonifractor plautii* (**Supplementary Tables 2 and 3)**.

Similar analyses were carried out for the Finnish FINRISK cohort, computing Cliff’s delta in a comparison of healthy individuals (n=2,525) with prevalent cancer cases (n=268). Of the 566 species identified by MetaPhlAn4 at ≥5% prevalence, 516 (91%) were matched to species of Meteor2 database, including 179 LKB species. Of these, 137 (76%) were enriched or depleted in the same direction as in the lung, kidney and breast cancers used to identify LKB species (**Supplementary Table 4**), a very strong bias towards coherence (*χ^2^ test* p=8.1E-13). Cliff’s delta between LKB and FINRISK cancers were also well correlated (Pearson R^2^= 0.14 to 0.33, **Supplementary Fig 1, G-K**). We conclude that cancer-related microbiome alterations are similar in different populations, irrespective of the geographical region in Europe, even if the magnitude of effect sizes vary, possibly due to population-specific dietary or genetic backgrounds.

To further test this conclusion, we compared Lifelines and FINRISK cohorts and identified 351 species present in both. Of these, 241 were coherently enriched or depleted in cancer cases relative to healthy controls, a very significant bias towards coherence (*χ^2^ test* p=2.7 E-12). Cliff’s delta values of all common species were positively correlated, as expected (Pearson R^2^ = 0.18, **Supplementary Fig 3 A)**. For 30 species differentially significant in abundance in both cohorts (Wilcoxon p<0.05) the correlation increased to R^2^ = 0.79; all were coherent (**Supplementary Fig 3 B**). These observations support our conclusion of common microbiome alterations in different cancers and different populations.

### Gut microbiome alterations associated with future cancer development

We hypothesized that people who will develop cancer in the future already have cancer-like alterations of their gut microbiome prior to their diagnosis. To test this hypothesis, we computed Cliff’s delta for species of LifeLines individuals with incident cancers (n= 245), in comparison to healthy controls (n=701), and found a high correlation with Cliff’s deltas computed for individuals with prevalent cancers (n=248) and the same healthy controls (Pearson R^2^= 0.39 for 524 species of the cohort, **Fig. 3A**). The strength of correlation increased to R^2^=0.59 for species significantly differentially abundant (p<0.05 by Wilcoxon test, n=173 species) in the comparison of prevalent cancers with healthy individuals (**Fig. 3B**). Of the 173 species, 155 (89.6%) were enriched (n=46) or depleted (n=109) in both prevalent and incident cancers, a very strong bias towards coherent species variation in the two groups (*χ^2^ test* p=2.1E-25). Correlation increased even further to R^2^=0.93 for species significantly differentially abundant in comparisons of prevalent and incident cancers with healthy individuals (n=39, **Fig. 3C**); all varied coherently. This indicates that alterations of the gut microbiome identified in prevalent cancers cases are already present in people who will develop cancer in the future. These alterations appear to be smaller in magnitude than in diagnosed cancer cases, as deduced from comparisons of absolute values of Cliff’s delta in prevalent and incident cancers, for both the cancer-enriched and cancer-depleted species (**Fig. 3D**, Student test p=5.6E-8 and p=2.7E-27, respectively).

**Fig 3.**
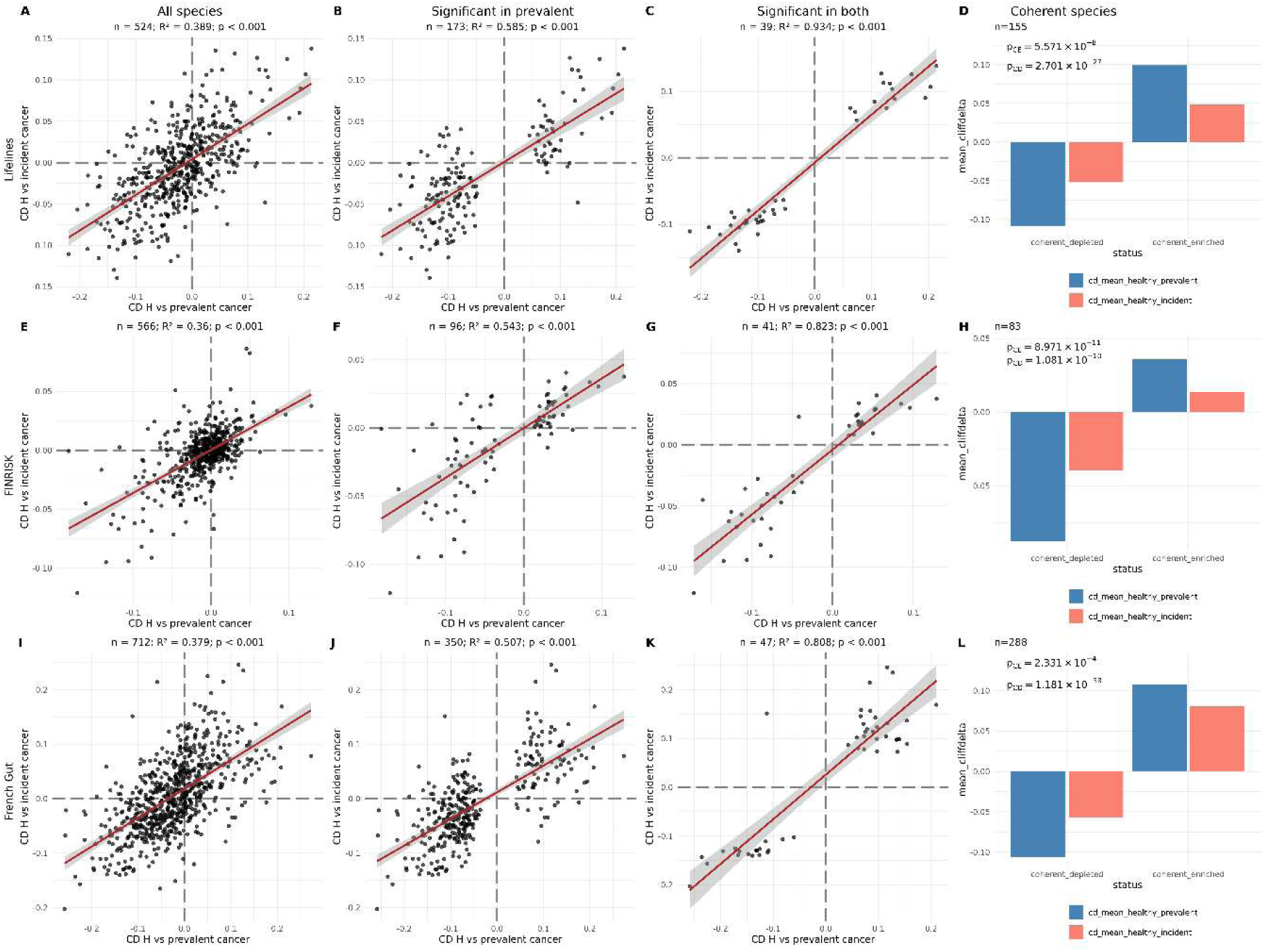
Coherence of gut microbiome alterations in prevalent and incident cancers. For the Lifelines cohort (A-D): Cancer cases (prevalent, n=248; incident, n=245) were compared with healthy controls (n=701), and Cliff’s delta values were computed for species common to the three groups for prevalent and incident cancers. Pairwise comparisons of Cliff’s deltas for all species with prevalence ≥10% (A), for species with significantly different abundances (Wilcoxon p<0.05) in healthy with prevalent cancer comparison (B) and for species with significantly different abundances (Wilcoxon p<0.05) in both comparisons (C); n indicates number of species for each comparison, p denotes significance of correlation, R^2^ denotes the squared Pearson correlation coefficient. The mean Cliff’s delta values of species that varied coherently in prevalent (at Wilcoxon p<0.05) and incident cancers are shown in panel D, negative and positive values correspond to species depleted and enriched in cancer, respectively; p values were computed using the two-tailed Student test, p_CD_ and p_CE_ refer to the p values for cancer depleted and cancer enriched species, respectively. Results of similar analyses are displayed for the FINRISK cohort (E-H; prevalent cancers n=268, incident cancers n=1,105, healthy controls n=2,525) and the French Gut cohort (I-L; prevalent cancers, n=463, incident cancers n=86, healthy controls, n=750).

Interestingly and importantly, the correlation of LKB species that varied coherently in prevalent cancers in Lifelines (n=128, **Supplementary Table 3**) was close (Pearson R^2^= 0.54, **Supplementary Fig. 4A**) to that of species significantly different between Lifelines prevalent cancers and healthy individuals (n=173, R^2^=0.59, **Fig. 3B**), illustrating again common cancer-related microbiome alterations. For LKB species with significant differences in abundance in prevalent cancers within the Lifelines cohort compared with healthy individuals (p<0.05 by Wilcoxon test, n=66), the correlation increased even further (R^2^= 0.67, **Supplementary Fig. 4B**).

Similar correlations of Cliff’s deltas between prevalent and incident cancers were observed in the FINRISK cohort, where prevalent and incident cancer cases (n=268 and n=1105, respectively) were compared with healthy controls (n=2525). The correlation strength increased from Pearson’s R^2^ = 0.36 for all species with ≥5% prevalence (n = 566) to R^2^ = 0.82 for significantly altered species (p<0.05 by Wilcoxon test) in both comparisons (n = 41, **Fig. 3E-G**). As in the Lifelines cohort, alterations of the microbiome in prevalent cancers were significantly higher than in incident cancers, as assessed by absolute Cliff’s delta values (**Fig. 3H**). Analysis of the French Gut cohort (prevalent cancers, n=463, incident cancers n=86, healthy controls, n=750, randomly selected from the total pool; the number chosen to match approximately Lifelines controls) yielded very similar results (**Fig. 3 I-L**). These analyses suggest that the microbiome may shift toward a cancer-associated composition well before cancer diagnosis.

To test this conclusion further, we examined the incident cancer cases (n=206) of the Latvian GISTAR cohort, by computing Cliff’s delta in comparison with healthy controls (n=251). As GISTAR only includes incident cases, we compared the incident cancer Cliff’s delta with the ones observed for LKB species in the French cohorts with diagnosed cancers. Correlations were positive for all the 3 diagnosed cancer types (lung, kidney and breast) with Pearson R^2^ between 0.32 and 0.56 (**Supplementary Fig 5**). Of the 275 LKB species 210 (76%) varied coherently in GISTAR (*χ^2^ test* p=9 E-21). We conclude that cancer-related microbiome alterations may be present in individuals who will develop cancer in the future. Whether these pre-diagnostic alterations might also reflect a possible presence of early, clinically occult malignancies, remains to be elucidated.

### Stratification of individuals into groups with differential cancer risk

We used the extent of alteration of the gut microbiome to stratify individuals into groups with different cancer risks. Alteration was assessed in four different ways, following the approach used for the stratification of individuals for the risk of Parkinson’s disease (Menozzi et al., 2026), by computing for each individual: (i) the relative abundance of species enriched in cancer, defined as the sum of abundances of enriched species divided by the sum of abundances of all species; (ii) the relative abundance of species depleted in cancer; (iii) the relative number of species enriched in cancer, defined as the number of cancer-enriched species divided by the number of all species, and (iv) the relative number of species depleted in cancer. The four measures are correlated but capture different aspects of microbiome composition – for instance, an individual could have higher abundance of fewer cancer-enriched species than another and their microbiome would appear to be more altered by the relative abundance score and less altered by the relative number score. The four scores were combined by ranking individuals for each score (ascending and descending ranks for cancer enriched and depleted species, respectively) and computing the average rank for each individual – the highest average ranks thus identify the most altered microbiomes. Analysis of the distribution of cancer cases average ranks in a cohort provides an estimate of the relative probability of having either a prevalent cancer or developing an incident cancer, which may function as a measure of cancer risk.

Stratification efficacy was first evaluated by pooling diagnosed cancer cases from the Oncobiotics (lung and kidney) and Canto (breast) cohorts with cancer-free individuals from the French Gut cohort (n = 5,115, representing the first chronological enrollees). Microbiome alterations were assessed as described in the previous paragraph, using the 275 LKB species. Each cancer type was stratified separately. All subjects - both cancer cases and healthy controls - were assigned to quintiles based on the distribution of their average ranks. The ratio of cancer cases assigned to the highest versus the lowest quintile (Q5/Q1) varied between 5.3 and 21.5 for Oncobiotics lung and kidney cancers, respectively, and was 5.7 for Canto (Supplementary Fig. 6A–C). This demonstrates that a greater extent of microbiome alteration correlates with a higher likelihood of having cancer.

Notably, these ratio values might be inflated because the stratified cancer cases were the same as those used to select the LKB species, even though substantially more controls were included in the stratification than in the selection step (ranging from a 9-fold increase for lung cancers to a 138-fold increase for kidney cancers). Consequently, this analysis could be considered as a training-set stratification. To address this limitation, we evaluated the Oncobiotics bladder cancer cases, which were not used for LKB species selection and thus served as an independent test set. For this cohort, the Q5/Q1 ratio was undefined because no cases were assigned to Q1; instead, 13 out of 14 cases fell into Q4 and Q5 (Supplementary Fig. 6D; exact binomial test p = 5.9E-5). Although limited by the small sample size of the bladder cancer subset, these results indicate that individuals with diagnosed cancers can be stratified simply by the extent of their microbiome alteration.

We next assessed the stratification efficiency of prevalent and incident cancer cases across the three population cohorts. Because information regarding microbiome variation in prevalent cancers was utilized to select LKB-coherent species, the stratification of prevalent cancers resembles a training-set analysis. In contrast, information from incident cancers was not used for species selection, making their stratification equivalent to an independent test-set analysis.

In the Lifelines cohort, utilizing 128 LKB-coherent species (**Supplementary Table 3; Fig. 4A, B**), the Q5/Q1 ratios between the extreme quintiles for prevalent and incident cancers were 3.5 and 1.9, respectively. These ratios increased slightly (to 3.7 and 2.0, respectively) when microbiome alterations were assessed using only the species that differed significantly in abundance between healthy controls and prevalent cancer cases (Wilcoxon *p* < 0.05, n = 66, data not shown). This demonstrates not only that Lifelines prevalent cancers can be stratified - albeit to a lesser extent than the Oncobiotics and Canto cohorts - but also, crucially, that incident cancers can be stratified as well.

**Fig 4.**
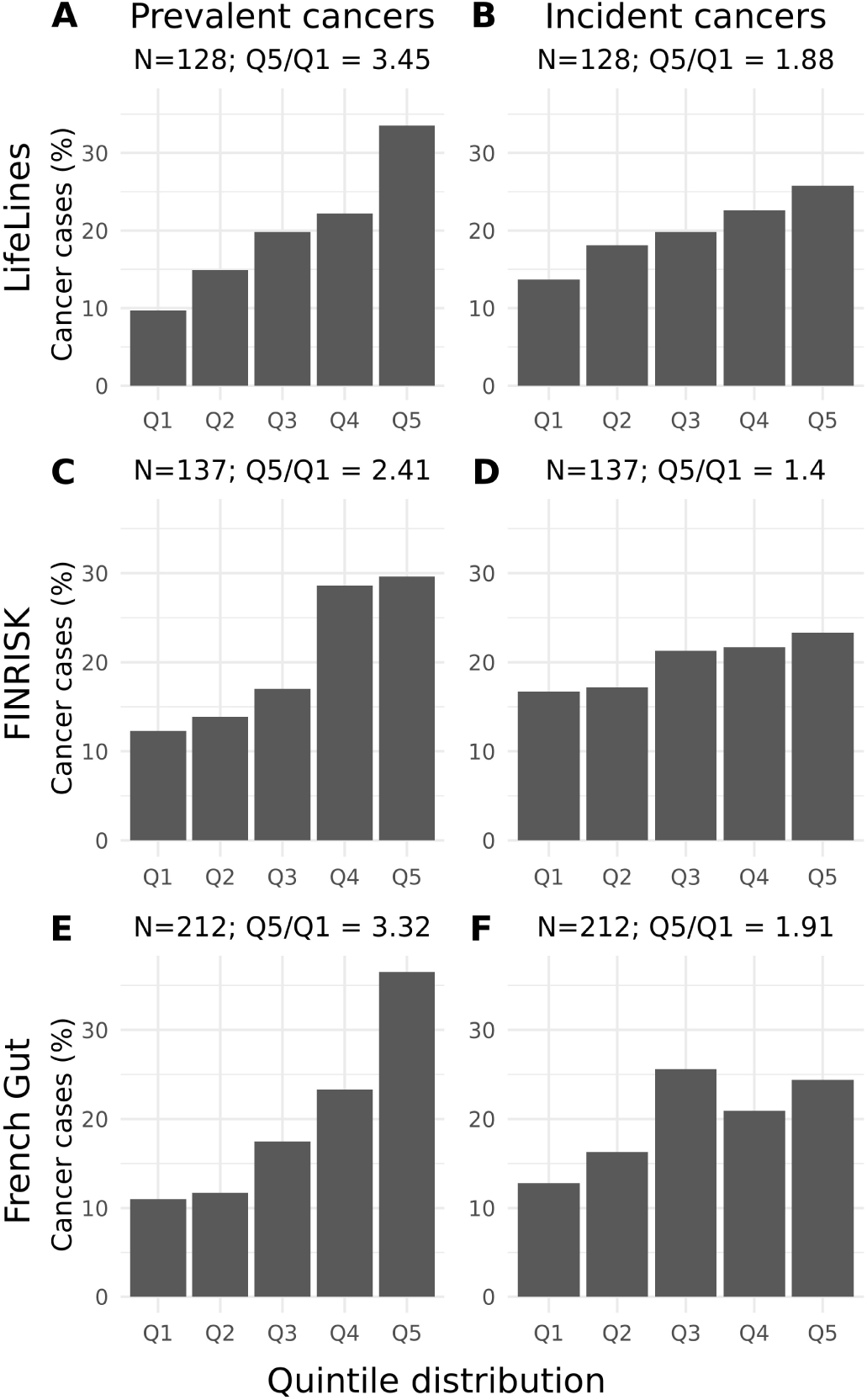
Stratification of prevalent and incident cancers by the level of gut microbiome alterations. For the Lifelines cohort, prevalent (A) and incident (B) cancers are presented. Each individual (controls, n=701) and either prevalent (n=248) or incident (n=245) cancer case was attributed an average alteration rank, and the individuals were placed in the quintiles of the rank distribution (Q1 to Q5). The proportion of cancer cases relative to total cancer cases in each quintile is displayed, the ratio between the extreme quintiles (Q5/Q1) is shown, N refers to the number of bacterial species used for stratification. Similar quintile distributions are displayed for the FINRISK cohort, prevalent (C) and incident (D) cancers; controls n=2525, prevalent cancers n=268, incident cancer n=1105 and for the French Gut cohort, prevalent (E) and incident (F) cancers; controls n=4,022, prevalent cancers n=463, incident cancers n=86.

A similar analysis of the FINRISK cohort using 137 LKB-coherent species (**Supplementary Table 4)** yielded comparable results for prevalent cancers (Q5/Q1 = 2.4, rising to 3.4 when restricting to the 41 species significantly different in abundance; data not shown) and a somewhat lower stratification efficiency for incident cancers (Q5/Q1 = 1.4; **Fig. 4C, D**). A comparable stratification pattern was observed in the French Gut cohort using 212 LKB-coherent species (**Supplementary Table 5; Fig. 4E, F**), with Q5/Q1 ratios of 3.3 and 1.9 for prevalent and incident cancers, respectively; no further improvement was observed when restricting to significantly different species (Wilcoxon *p* < 0.05, n = 161). Notably, many more controls (5.4-fold; n=4022, all individuals matching the inclusion criteria) were used for stratification than for Cliff’s delta analysis (n=750). Finally, GISTAR incident cancers were also effectively stratified using the 275 LKB species (Q5/Q1 = 1.6, **Supplementary Fig. 7**). Taken together, these results suggest that cancer-related alterations of the gut microbiome can stratify individuals without a cancer diagnosis into groups with higher or lower probabilities of developing cancer in the future.

## Discussion

The capacity of a deviant gut microbiome to influence a chronic inflammatory disorder as complex as cancer development has remained an open question. Conversely, it can be envisaged that individuals with life conditions favoring the development of a deviant intestinal microbiome may be at inherently higher risk of cancer. Research investigating the clinical significance of gut dysbiosis in cancer patients has focused on cancer prognosis (or staging at diagnosis) as well as prediction of treatment response, toxicity and long-term survival. In the present study, we addressed the possibility of assessing cancer risk in undiagnosed individuals by the level of gut microbiome alteration.

To this goal, we deployed a Cliff’s delta-based approach of microbiome characterization, adapting a framework originally designed to address the risk of Parkinson’s disease among disease-free individuals (Menozzi et al., 2026). We report coherent microbiome alterations in different types of cancers (lung, kidney, breast and bladder) relative to cancer-free individuals matched by age, BMI, and sex; antibiotic treatments and smoking did not confound our findings. The 275 species involved, termed LKB species, represent about 40% of the microbiome in both cancer cases and healthy individuals. Previous studies, focusing on species with statistically significant differences in abundance between individuals with and without cancer, already reported many of these species as cancer-enriched or depleted, in alignment with our observations (Thomas et al., 2023) for recent meta-analysis); however, the large extent of cancer-related microbiome alterations we describe was not captured. By evaluating coordinated ensembles of species rather than individual taxa, our framework significantly enhances statistical power to uncover extensive, disease-related microbiome alterations..

We found common cancer-related microbiome alterations in three large population studies, Lifelines, FINRISK and French Gut. Comparison of cancer cases diagnosed prior to microbiome sampling with unmatched healthy controls (BMI below 30, non-smokers, free of chronic diseases) revealed that a large majority of LKB species (76-80%) varied in the same direction as in comparisons of cancer cases matched to controls by BMI, gender and age and maintained a high topological invariance (R² up to 0.64). This robust preservation of the species-ensemble hierarchy suggests that absence of matching does not preclude detection of cancer-related microbiome alterations and indicates that our analytical framework tracks a genuine, unconfounded biological signal.

Remarkably, the coherent cancer-related microbiome alterations identified here were detected despite significant methodological heterogeneity across cohorts. These variations span sample collection (five protocols, including FIT buffer residues), DNA extraction (five protocols), sequencing (three platforms: Ion Torrent, Illumina, and MGI), and bioinformatic analysis (two suites: Meteor 2 and MetaPhlAn4). This consistency underscores the robustness and real-world utility of our Cliff’s delta-based approach, especially where methodologies cannot be standardized or retrospectively aligned—as is typical in large-scale prospective studies like Lifelines and FINRISK. The resilience of the approach is likely attributable to the use of intra-cohort analyses, where experimental methodologies are consistent between cases and controls. Nevertheless, adopting standard protocols across cohorts whenever feasible would undoubtedly refine similar microbiome analyses. For instance, the detection and quantification of critical, low-abundance cancer-enriched species may require both sufficient sequencing depth and specialized tools. In this context, platforms like Meteor2 are particularly effective at capturing low-abundance species across diverse cohorts (Ghozlane et al., 2024).

Importantly, the repurposing of FIT buffer residues for microbiome analysis—and the demonstrated consistency of these results with standard collection methods—presents a highly promising avenue. With hundreds of millions of FIT samples generated annually through global screening mandates, this approach creates an unprecedented opportunity for large-scale, population-wide microbiome monitoring within the critical age window for cancer risk acceleration.

Yet another appealing facet of the Cliff’s delta-based approach is that it obviates the need to pool the data in a single central facility prior to analysis. Such centralization is becoming increasingly difficult due to the privacy protection rules limiting the release of potentially identifying information. Indeed, no primary data of the three large population cohorts, Lifelines, FINRISK and French Gut were shared in our study – only algorithms and outcomes of analyses were exchanged, aligning with federated learning strategies currently emerging in the microbiome field (Späth et al., 2024). More generally, distributed population cohorts provide a natural setting for learning modality-specific representations across diverse populations while keeping cancer-specific adaptation downstream.

Beyond coherence and extent of microbiome changes in different cancer types, Cliff’s delta-based analyses revealed similar microbiome alterations in individuals with cancers diagnosed prior to microbiome sampling (prevalent cancers, n=979) and individuals who developed cancers after sampling (incident cancers, n=1,436) in three independent studies from three different countries. The fourth cohort, from Latvia included only incident cancers (n=206), with alteration similar to those in French cancer cases. Our findings thus indicate that cancer-related microbiome alterations may take place ahead of cancer development. Expectedly, the extent of alteration was smaller in individuals at risk of cancer than in individuals with cancer, as inferred from the Cliff’s delta magnitude.

Stratification of people into groups at higher and lower risk of cancer would create opportunities to mitigate cancer risk. We show here that such stratification might be possible, simply by assessing the level of cancer-related microbiome alteration. Considering that people that developed cancers after microbiome sampling were at high risk of cancer at the time of sampling, we show that the individuals belonging to the extreme quintiles of the distribution of microbiome alterations differ by up to twofold in the relative risk of cancer.

An obvious limitation of our study is its reliance on a single-time-point analysis. Longitudinal studies would likely be more informative, as unrecorded lifestyle changes could alter the microbiome composition of individuals who remain cancer-free despite exhibiting microbial alterations at the time of sampling. Conversely, individuals with minimal microbiome alterations at baseline might shift toward a cancer-associated microbial profile due to subsequent lifestyle changes. Consequently, regular microbiome monitoring may improve cancer risk stratification.

Furthermore, we cannot exclude the possibility that some individuals in the control group had undiagnosed, subclinical cancers at the time of sampling. Exploring the relationship between the magnitude of microbiome alteration and the time to cancer diagnosis, in conjunction with longitudinal monitoring, could help differentiate the early detection of existing malignancies from the assessment of future cancer risk *per se*, both of which hold substantial clinical value and define distinct targets for downstream cancer-specific modelling.

From a clinical translation perspective, a third limitation is our reliance on high-throughput sequencing of the entire microbiome rather than targeting a selected panel of biomarkers. However, large population cohort studies have already overcome several associated hurdles by establishing simple, standardized sample collection protocols, reducing sequencing costs and turnaround times, and streamlining bioinformatic pipelines. These technological advances will likely facilitate widespread microbiome monitoring in the future - potentially comparable to routine blood tests - offering a powerful tool to identify the risk of deadly diseases such as cancer.

Additionally, our approach, which relies on effect-size comparisons, is inherently susceptible to residual confounding. Several cancer-associated LKB species are known to correlate with lifestyle factors and medication usage, which are likely to differ between cancer cases and healthy controls. Although we demonstrated that the most common confounders, such as smoking and antibiotic use, do not significantly impact the detection of cancer-related species, other unmeasured factors may still contribute. Nonetheless, the microbiome may capture a composite signal of exposures and host states associated with oncogenesis. Such a tool could be broadly integrated into existing cancer screening frameworks (Birkeland et al., 2026; Byrd et al., 2026; Louca et al., 2026), offering substantial clinical utility for early cancer detection and risk prediction.

Finally, while the LKB-species-derived signature, largely trained on lung cancer cases, demonstrated generalizability across diverse cancer types, it remains unclear how well it extends to all forms of neoplasia. Although common cancers are well-represented in our cohorts, our dataset includes fewer colorectal and pancreatic cancers and lacks inherited malignancies that develop in the context of local inflammation or germline mutations. External validation across these settings will allow to determine which components of the microbiome representation are general and which might require cancer-specific adaptation.

Irrespective of these limitations, stratification of individuals without a cancer diagnosis at sampling provides a proof-of-concept framework for microbiome-based cancer risk estimation. An important next step is to test whether microbiome-derived representations add predictive value beyond established epidemiological and clinical risk factors. Such modality-specific representations, learned across heterogeneous populations, could subsequently be integrated with clinical and other molecular modalities for cancer-specific risk stratification. This separation between population-scale representation learning and cancer-specific multimodal integration provides a natural path toward foundation-model strategies for risk prediction.

## Acknowledgements

MF is supported by the SEERAVE Foundation and the European Union’s Horizon Europe research and innovation program under grant agreement No 101095604 [project acronym: PREVALUNG-EU, project title: Personalized lung cancer risk assessment leading to stratified Interception]. GV was supported by Oncobiotics grant ANR-21-RHUS-0017. LG was supported by PEPR1071 SAMS PREANALYTICS grant ANR-24-PESA-0004. Le French Gut was supported by MetaGenoPolis grant (ANR-11-640 DPBS-0001), PEPR SAMS PC Cohortes-Microbiomes (ANR-24-PESA-0005), Carnot Institute Qualiment (#20 CARN 0026 01), Genoscope, the Commissariat à l’Energie Atomique et aux Energies Alternatives (CEA), France Genomique (ANR-10-INBS-09-08), APHP and Fondation Carnot APHP. MA, RB, ELC and YR were additionally supported by the European Union’s Horizon research and innovation programme Microb-AI-ome project under the Grant Agreement n° 101079777. Views and opinions expressed are those of the author(s) only and donot necessarily reflect those of the European Union. RKW is supported by the Seerave Foundation, the Innovation Health Initiative Joint Undertaking (IHI JU) grant INTERCEPT (101194780), and the EU Horizon Health consortium grant ID-DarkMatter-NCD (101136582).

## Author contributions

Conceptualization: SDE, LZ; Methodology: SDE, JB, GV, JRB; Software: MA, GV, ELC, LG, JB, YR; Validation: SDE, MA, GV, YR; Formal analysis: SDE, GV, LG; Resources: MA, JK, JRB, RKW, ML, OK, LL, TN, RB, LD, JR; Data curation: MA, MF, ELC, GV, YR, LG, LD; Writing – original draft: SDE, GK, LZ, MF, GV, MA; Revision: EB, JRB, RKW, TBR, GK, BR, BP, AE, LD, NP. Visualization: SDE, GV, LG; Supervision: SDE, LZ, MA, JB; Project administration: SDE, LZ, MA; Funding acquisition: LZ, MA, SDE, JB, PV.

## Data availability

Oncobiotics: Metagenomic data from the retrospective Oncobiotics cohort are publicly available through the European Nucleotide Archive (ENA; PRJEB22863, https://www.ebi.ac.uk/ena/browser/view/PRJEB22863). Prospective data are deposited under PRJEB81798 (https://www.ebi.ac.uk/ena/browser/view/PRJEB81798) and are currently under embargo and will be released upon publication.

Canto: Raw data for the metagenomes newly sequenced in this study are available in NCBI-SRA under the BioProject PRJNA718520.

Lifelines: All metagenomes analysed in this study will be made available upon publication. Metadata can be requested directly through Lifelines.

FINRISK: The metagenomic data are available from the European Genome-Phenome Archive (accession number EGAD00001007035). The phenotype data contain sensitive information from healthcare registers and they are available through the THL biobank upon submission of a research plan and signing a data transfer agreement (https://thl.fi/en/web/thl-biobank/for-researchers/application-process).

Le French Gut: To protect participant privacy and comply with the French Data protection authority (CNIL) authorization granted for Le French Gut study, individual-level metagenomic data cannot be made openly available without access control. Metagenomic data will be deposited in the European Genome-phenome Archive (EGA) under a specific EGA accession number. Data can be made available upon request through the EGA data access mechanism. All requests will be reviewed by the Data Access Committee of Le French Gut study. Proposals, researchers or institutions requesting data will be approved if they meet the standard criteria related to ethics, privacy and data protection regulations. If the collaboration is approved, a data access agreement will be required, and any necessary authorizations from the relevant administrative authorities may be needed. In compliance with existing regulations, no personally identifiable data, as well as SNDS data, will be accessible.

GISTAR: this is an ongoing study as study subjects are continuously followed. Therefore, individual participant metagenomic and clinical data are not publicly available and cannot be deposited in public repositories. The availability of either metagenomic or other participant data is a matter of decision of the Data Safety and Monitoring Board. The requests should be addressed to the following e-mail address.

## Code availability

The R code developed for the analyses performed on both Le French Gut and the French cancer cohorts is available at https://forge.inrae.fr/french-gut/fg_cancer.

## Le French Gut Consortium

Najate Achamrah¹,²,³, Mathieu Almeida⁴, Anne-Sophie Alvarez⁴, Mourad Benallaoua⁵, Robert Benamouzig⁵, Magali Berland⁴, Oana Bernard⁶, Sylvie Binda⁷, Anne Blais⁵, Hervé Blottière⁴,⁸, Elise Borezée-Durant⁴, Léa Breton⁹, Alexandre Cavezza⁴, Benoît Chassaing¹⁰, Lison Chevreau⁴, Moïse Coéffier¹, Chloé Connan⁴, Anne-Marie Davila-Gay¹¹, Lauren Demerville¹², Sébastien Dime⁴, Kahina Djerrah⁵, Michel Dojat¹³, Joël Doré⁴,¹⁴, Assia Dreux¹⁵, Inès Drouard⁴, Franz Durandet³⁷, Olivier Durlach¹⁶, Erik Eckhardt¹⁷, Alexandre Famechon⁴, Etienne Formstecher¹⁸, Clémence Frioux⁴,¹⁹, Sébastien Fromentin⁴, Nathalie Galleron⁴, Rozenn Gazan²⁰, Amine Ghozlane²¹, Marine Gilles⁴, Oscar Gitton-Quent⁴, Lindsay Goulet⁴, Anne Hiol⁴, Esra Ilhan¹⁴, Evelyne Jouvin-Marche²³, Simon Labarthe²⁴, Nicolas Lapaque⁴,¹⁴, Milan Lazarevic¹², Emmanuelle Le Chatelier⁴, Julie Lê-Hoang⁴, Marion Leclerc⁴,²⁵, Patricia Lepage¹⁴, Emmanuelle Maguin¹⁴, Matthieu Maillot²⁰, Claudine Manach²⁶, Emile Mardoc⁴, Mahendra Mariadassou²⁷, Elliot Mathieu⁴, Nicolas Maziers⁴,²⁸, Idir Mazouzi⁵, Juliette Meyer²¹, Bénédicte Monnerie⁴, Christian Morabito⁴, Christine Morand²⁶, Julie-Anne Nazarre²⁹, Sophie Nicklaus³⁰, Anne-Sophie Nyob³¹, Pedro H. Oliveira²², Florian Plaza Oñate⁴, Nicolas Pons⁴, Benoît Quinquis⁴, Xavier Raffoux⁴, Etienne Ruppé³²,³³, Anne Rutigliano³¹, Jean-Marc Sabaté⁵, Jacques Sainte-Marie³⁴, Mathilde Sola⁴,¹⁹, Guilhem Sommeria-Klein²⁴, Manon Sudrie⁴, Julien Tap¹⁴, Florence Thirion⁴, Vincent Thomas³⁵, Patrick Trieu-Cuot³⁶, Karine Valeille⁴, Charline Vasseur⁴, Patrick Veiga⁴,¹⁴, Florent Vieux²⁰, Giacomo Vitali⁴, Fabien Wuestenberghs⁵.

¹ Univ. Rouen Normandie, Inserm, ADEN UMR1073, CHU Rouen, CIC-CRB 1404, Department of Nutrition, F-76000, Rouen, France; ² Univ. Rouen Normandie, Institute for Research and Innovation in Biomedicine (IRIB), 76000, Rouen, France; ³ Department of Nutrition, CHU Rouen, 76000, Rouen, France; ⁴ Université Paris-Saclay, INRAE, MetaGenoPolis, 78350, Jouy-en-Josas, France; ⁵ Department of Gastroenterology, Avicenne Hospital, APHP, Université Paris Nord - La Sorbonne, Bobigny, France; ⁶ Biocodex, Gentilly, France; ⁷ Rosell Institute for Microbiome and Probiotics, Montréal, Québec, H4P 2R2, Canada; ⁸ Nantes Université, INRAE, UMR1280, PhAN, Nantes, France; ⁹ INSERM, Paris, France; ¹⁰ Microbiome-Host Interactions, INSERM U1306, CNRS UMR6047, Institut Pasteur, Université Paris Cité, Paris, France; ¹¹ Université Paris-Saclay, AgroParisTech, INRAE, UMR PNCA, Palaiseau, France; ¹² APHP, Paris, France; ¹³ Université Grenoble Alpes, Inserm U1216, Grenoble Institut Neurosciences, Inria, Grenoble, France; ¹⁴ Université Paris-Saclay, INRAE, MICALIS, 78350, Jouy-en-Josas, France; ¹⁵ Greentech, Saint-Beauzire, France; ¹⁶ Institut du Vieillissement, Hospices Civils de Lyon, Lyon, Auvergne-Rhône-Alpes, France; ¹⁷ DSM-Firmenich, Houdan, France; ¹⁸ GMT Science, Paris, France; ¹⁹ Inria, University of Bordeaux, INRAE, 33400 Talence, France; ²⁰ MS-Nutrition, Marseille, France; ²¹ Institut Pasteur, Université Paris Cité, Bioinformatics and Biostatistics Hub, F714 75015, Paris, France; ²² Génomique Métabolique, Genoscope, Institut François Jacob, CEA, CNRS, Université d’Evry, Université Paris-Saclay, Evry, France; ²³ Institut Physiopathologie, Métabolisme, Nutrition (PMN) Inserm, Paris, France; ²⁴ INRAE, BIOGECO, Univ. Bordeaux, Cestas, France; ²⁵ Université Clermont Auvergne, INRAE, MEDIS, Clermont, France; ²⁶ Université Clermont Auvergne, INRAE, UNH, F-63000 Clermont-Ferrand, France; ²⁷ Université Paris-Saclay, INRAE, MaIAGE, 78350, Jouy-en-Josas, France; ²⁸ Hospital Center Sud Francilien, Intensive Care Unit, Corbeil-Essonnes, France; ²⁹ CarMeN Laboratory, Université Claude Bernard Lyon 1, INSERM, INRAE, Pierre Bénite, France; ³⁰ INRAE, Paris, France; ³¹ ANJAC, Paris, France; ³² Université Paris Cité and Université Sorbonne Paris Nord, Inserm, IAME, F-75018 Paris, France; ³³ AP-HP, Hôpital Bichat-Claude Bernard, Laboratoire de Bactériologie, F-75018 Paris, France; ³⁴ Inria Paris, Team Ange, Paris, France; ³⁵ Danone, Gif-sur-Yvette, France; ³⁶ Institut Pasteur, Université Paris Cité, Unité de Biologie des Bactéries Pathogènes à Gram-positif, Paris, France; ³⁷ IAGE, Montpellier, France.

## Methods

### Cohorts

The two cohorts of cancer cases, Oncobiotics (NCT04567446) (Derosa et al. 2022) and CANTO (NCT01993498) (Terrisse et al., 2021) have been previously described, as were the four population-based cohorts, LifeLines, FINRISK (Salosensaari et al., 2021), Le French Gut (NCT05758961) (Connan et al., 2025) and GISTAR (Leja et al., 2017). Relevant information is summarized in Table 1 and Supplementary Table 1. Translational analyses were performed under the study ONCOBIOTICS (sponsor protocol no. CSET 2017/2619, ID-RCB no. 2017-A02010-53) according to the ethical guidelines and approval of the local ethical committee (CPP Est-III, Kremlin-Bicêtre Hospital). Translational analyses were performed within the framework of the CANTO cohort study (promoted by UNICANCER, protocol no. UC-0140/1103, CSET 2012/18291, national identifier ID-RCB no. 2011-A01095-36). The study protocol was approved by the French regulatory authorities (Afssaps, ref: B111158-20) and the French ethics committee (CPP / Institutional Review Board, approval no. 11-039) on October 14, 2011. The trial is registered on ClinicalTrials.gov under identifier NCT01993498.

### Lifelines cancers and controls

Among participants in the Lifelines Biobank, 5,997 adults with stool metagenomic data generated within the Dutch Microbiome Project (DMP) were included in this analysis. The Lifelines protocol was approved by the Medical Ethical Committee of the University Medical Center Groningen (METc 2017/152), and DMP participants provided additional written informed consent.

Cancer diagnoses were ascertained through linkage to the nationwide Dutch Pathology Database (PALGA), with records available through 16 March 2023. PALGA provides histopathological confirmation of malignancy and detailed tumor characteristics, including histological subtype, anatomical site, tumor size, and stage. Malignancies were categorized according to the Netherlands Comprehensive Cancer Organisation (IKNL) classification. Cancer diagnoses occurring before stool sampling, premalignant or benign lesions, uncertain cancer diagnoses and self-reported previous cancer without a corresponding PALGA record were excluded

Incident cancer was defined as the first histopathologically confirmed malignancy recorded in PALGA after stool collection. Participants were followed from stool collection, conducted between January 2013 and September 2016, until their first cancer diagnosis or the end of PALGA follow-up on 16 March 2023, providing up to approximately 10 years of follow-up. During this period, 245 participants were diagnosed with incident cancer, whereas 5,752 remained cancer-free.

### FINRISK cancers and controls

The study protocol of FINRISK 2002 was approved by the Coordinating Ethical Committee of the Helsinki and Uusimaa Hospital District (Ref. 558/E3/2001). All participants signed an informed consent. In FINRISK, 268 participants had diagnosed cancer prior to the sample collection. Cancer diagnoses were confirmed by linking the records to the nationwide Finnish Cancer Registry using unique personal identity codes, with records available until 2024. Notification of cancer to the Finnish Cancer Registry has been mandatory since 1953, and its coverage of solid tumours is essentially complete (>99%), with diagnoses confirmed histopathologically in the large majority of cases. The registry records topography and morphology coded according to ICD-O-3, together with the date and basis of diagnosis and the extent of disease at diagnosis (local, regional, distant). Incident cancer was defined as the first registry-confirmed malignancy recorded after stool collection. Participants were followed from stool collection in 2002 until their first cancer diagnosis, death, or the end of registry follow-up on 2024, providing up to approximately 22 years of follow-up. Death cases were obtained through linkage to the Statistics Finland Causes of Death Register. During this period, 1105 participants were diagnosed with incident cancer, whereas 5858 remained cancer-free. The healthy control group was defined as those with BMI<=30, no smoking, and no diagnosed cancer before or during the study period.

### French Gut cancers and controls

Le French Gut study has been approved by the the “Comité de Protection des Personnes” Sud-Est IV (CPP, 21.00225.000006) and the “Commission Nationale Informatique et Liberté” (CNIL, DR-2022-141). Within the French Gut project cohort, prevalent cancer cases were identified using a dual approach. First, cases were retrieved from the Système National des Données de Santé (SNDS)—the French National Health Data System— based on the registration of an Affection de Longue Durée (ALD 30) status, which denotes a medically certified and legally recognized long-term illness for malignant tumors. Second, cases were detected via the self-reported questionnaire administered to participants at baseline inclusion. Among the 463 identified cancer cases, 413 were detected through the SNDS, including 187 cases that were concurrently captured by the questionnaire. Conversely, 50 cases were identified exclusively through the questionnaire and lacked an official ALD 30 status. Incident cancer cases (n=86) were identified exclusively through the SNDS.

### GISTAR cancers and controls

For GISTAR the Ethics Committee of IARC has approved the study protocol 26/03/2013 and the relevant protocol updates 02/10/2015. reg. No. IEC 12–36; the Ethics Committee of Riga East University Hospital Support foundation has approved the protocol 03/10/2013, reg. No. 14-A/13, and the Central Medical Ethics Committee in Latvia has approved the protocol 09/12/2013, reg. No. 01–29.1/11. Average risk population sample from Latvia was recruited to the GISTAR cohort between 2013 and 2023 (Leja et al., 2017). Following the recruitment, the individuals were followed up for incident cancer diagnosis having been reported to the National Cancer Registry.

### Samples collection and processing

#### Cancer cohorts

Stool samples were self-collected by the participants at home according to the IHMS_SOP 004 (https://human-microbiome.org/index.php?id=SopCnum=004). Immediately after the collection, they were stored at −20 °C and transported on dry ice. Upon arriving at the biobank of the recruitment center (between 4 and 24 h later), they were stored at −80°C in plastic tubes. All the samples were processed according to International Human Microbiome Standards (IHMS) guidelines (SOP 03 V1). DNA was extracted from an aliquot of the stool samples using the IHMS SOP 07 V2 H and sequenced with an Ion proton sequencer (Thermo Fisher Scientific). The sequencing was performed in one single site (MetaGenoPolis).

#### Population cohorts

##### Le French Gut

Stool samples were collected and processed as described by Connan et al (Connan et al., 2025); DNA was purified from sample aliquots using the publicly available Epsilon pipeline (https://forge.inrae.fr/metagenopolis/epsilon-pipeline/).

Sequencing was performed using DNBSEQ technology (MGI Tech). Prior to sequencing, DNA degradation was checked using the DNA genomic 50kb kit on a Fragment Analyzer 5200 (Agilent) (Quinquis et al., 2024). After fragmentation of 100 to 500 ng of DNA, libraries were constructed according to the instructions in the MGIEasy Universal DNA Library Prep Set, FS or Fast FS Library Prep Set (MGI), including end repair, A-tailing, adapter ligation, and selection of constructs by amplification. Barcoded libraries were circularized using the MGIEASY Circularization Kit. Quality controls were performed using Small Fragment kits on a Fragment Analyzer 5200 (Agilent Technologies), Quant-iT dsDNA and Qubit ssDNA Assay Kits (Thermo Fisher Scientific). Circularized library pools are constructed to optimize the yield of 20 million paired-end reads (2 x 150) on a DNBSEQ G400 sequencer (MGI). Most steps were automated using Beckman Coulter and MGI pipetting handlers.

Remaining DNA and sample aliquots were stored at -70 °C in the MetaGenoPolis-SAMBO BRC (Biological Resources Center), part of INRAE (French National Research Institute for Agriculture, Food, and the Environment).

##### Lifelines

Stool samples were collected and processed as described by Gacesa and colleagues (Gacesa et al., 2022). In short, stool samples were collected between January 2013 and 2016. Participants collected stool samples at home and froze them within 15 minutes of defecation. Frozen samples were collected by LifeLines, transported on dry ice, and stored at -80°C at the University Medical Center Groningen (UMCG). DNA was isolated using the QIAamp Fast DNA Stool Mini Kit (Qiagen) on the QIAcube automated sample preparation system (Qiagen). Samples with DNA yields below 200 ng (measured using a Qubit 4 Fluorometer) were prepared using the NEBNext Ultra DNA Library Prep Kit for Illumina, whereas all other samples were prepared using the NEBNext Ultra II DNA Library Prep Kit for Illumina. Metagenomic sequencing was performed by Novogene on the Illumina HiSeq 2000 platform, generating approximately 8 Gb of 150 base pair paired-end reads per sample.

##### FINRISK

Stool samples were collected in the FINRISK 2002 population survey, a stratified random sample of adults aged 25–74 years drawn from six geographical regions of Finland (Borodulin et al., 2018). At the baseline examination, willing participants received a sampling kit with detailed instructions and collected the sample at home. Samples were mailed overnight under Finnish winter conditions to the laboratory of the Finnish Institute for Health and Welfare, where they were stored at −20 °C, and were transferred frozen in 2017 to the University of California San Diego for sequencing. Shotgun metagenomic sequencing was successfully performed for 7,231 participants on an Illumina HiSeq 4000 instrument (Salosensaari et al., 2021).

##### GISTAR

Sample collection tubes intended for the use of faecal immunochemical occult blood testing OC-Sensor from Eiken Chemical Co., Ltd (Tokyo, Japan) were used for microbiome sample collection, transporting and storage, and have been shown to be reliable for large-scale gut microbiome profiling (Gudra et al., 2019). All the returned sample tubes were immediately frozen at -80°C after a quantitative immunochemical faecal occult blood test was performed and were stored and transported frozen until further processing for microbiome analysis. Sample processing took place within 60 days from sampling.

Before the extraction, the total buffer content containing faecal material (∼1,5 ml) was extracted from the sample bottles with a sterile, single-use syringe and concentrated by lyophilization. Afterwards, microbial DNA was extracted from a lyophilized sample using FastDNA Spin Kit for Soil (MP Biomedicals, USA) and FastPrep 24 5G instrument. Additionally, DNA was extracted from a negative control and microbial community and DNA standards (ZymoBIOMICS, USA). Sequencing was performed by MGI-Tech Latvia laboratories, where the extracted DNA was fragmented in approximately 400 bp fragments using a Covaris S220 and a sequencing library prepared using MGIEasy PCR-Free DNA kit (MGI-tech, China) following the manufacturer’s standard protocol. Library quality was checked with Agilent Bioanalyser 2100 (Agilent Technologies, USA). Shotgun metagenomic sequencing was performed on the DNBSEQ-G400 platform (MGI, China) with 150bp paired-end reads.

### Filtering, cleaning and host reads removal

#### Cancer cohorts

##### French Gut and GISTAR

DNA reads were quality trimmed and filtered from sequencing adapters using fastp (v0.23.2) with the following parameters: *“--cut_front –cut_tail –n_base_limit 0 -- length_required C0 --trim_poly_g --adapter_sequence AAGTCGGAGGCCAAGCGGTCTTAGGAAGACAA –adapter_sequence_r2 AAGTCGGATCGTAGCCATGTCGTTCTGTGAGCCAAGGAGTTG”*. For GISTAR, we used the same fastp parameters except --n_base_limit changed to 3, to reduce the number of discarded reads in some samples. The remaining human DNA was removed using bowtie2 (v2.5.1) to align the reads to the human reference genome T2T-CHM13v2.0, using at least 90 % nucleotide identity threshold for filtering, employing samtools (v1.9) with the parameters “-f 4 -F 256 –N”.

#### Lifelines

Illumina adapters and low-quality bases were trimmed with Trimmomatic as implemented in KneadData v0.12.4 (http://huttenhower.sph.harvard.edu/kneaddata), using a leading and trailing base-quality threshold of Phred 20 and a 4-base sliding window requiring mean Phred ≥ 20. Reads shorter than 50 bp after trimming were discarded. Reads mapping to the human reference genome (GRCh37/hg19) were removed with Bowtie2 v2.5.1 (Langmead & Salzberg, 2012) in --very-sensitive end-to-end mode, with any aligning read discarded. Read quality was assessed with FastQC v0.12.1 before and after processing.

#### FINRISK

Reads were trimmed for quality and adapters with Atropos, and host reads were removed by mapping against the human genome assembly GRCh38 with Bowtie2.

### Taxonomic profiling

#### Cancer cohorts, French Gut and GISTAR

Filtered reads were separately mapped with METEOR v2 (v2.0.22) (Ghozlane et al., 2024) against a gut and oral catalogues comprising 10.4M (Wen et al., 2017) and 8.4M (Le Chatelier et al., 2023) reference genes respectively, and the resulting profiles were merged. The two catalogs were previously organized into 1,990 and 853 Metagenomic Species Pangenome (MSP) species (Le Chatelier et al., 2021; Nielsen et al., 2014; Plaza Oñate et al., 2019; Plaza Oñate & Le Chatelier, 2020) that correspond to clusters of co-abundant genes used as proxies for microbial species and containing core and accessory genes. MSP definition and taxonomy are available from Data INRAE ((Le Chatelier et al., 2021) and (Plaza Onate et al., 2021)). Relative abundance of a given MSP was computed as the mean abundance of its 100 ‘marker’ genes (that is, the genes that correlate the most altogether). If fewer than 10% of ‘marker’ genes were detected in a sample, the abundance of the MSP was set to 0. MSP richness was computed as the number of detected MSP species in a particular sample. All samples were checked for potential cross-contamination using the Meteor2 MSP abundance with CroCodeEL (v1.0.8) (Goulet et al., 2026). Samples were excluded if species added by contamination exceeded 12% or if the contamination rate was > 1 % with additional species exceeding 10%. Remaining suspicious samples were then flagged using CroCodeEL interpreter (https://metagenopolis.github.io/CroCoDeEL_interpreter) and discarded from the study.

#### Lifelines

Taxonomic profiles were generated using MetaPhlAn4 with the mpa_vOct22_CHOCOPhlAnSGB_202212 database and default parameters.

#### FINRISK

Filtered reads were processed with MetaPhlAn (4.1.1) using the default database version (mpa_vJun23_CHOCOPhlAnSGB_202403) for taxonomic mapping.

### Computational analysis

The comparison of species between groups in the cancer cohorts, French Gut and GISTAR cohorts was performed using the testRelation2 function from the momr package V1.3161. This function generates a matrix that provides statistical results calculated with the two-sided Wilcoxon rank-sum test, with the Benjamini-Hochberg procedure for multiple testing, and quantifies the magnitude of these differences using Cliff’s Delta (CD) for each species.

Coherent and non-coherent enriched or depleted species were identified based on the testRelation2 output, using the direction of CD as a guide. A dedicated R-script was created for LifeLines and FINRISK cohort – its output was compared with that of testRelation 2 and found to be identical for the same parameters.

Metagenomic species of Meteor and SGBs of MetaPhlAn were related via taxonomic annotation, using GTDB_r226_taxonomy and the chocophlan_mpa_Jun23 database.

Microbiome alterations were assessed as in Menozzi et al., 2026, separately for each cohort, by computing for every individual, controls and cancer cases: (i) relative abundance of species enriched in cancer, defined as the sum of abundances of enriched species divided by the sum of abundances of all species; (ii) relative abundance of species depleted in cancer; (iii) relative number of species enriched in cancer, defined as the number of cancer-enriched species divided by the number of all species, and (iv) relative number of species depleted in cancer. The four scores were combined by ranking individuals for each score (ascending and descending ranks for cancer enriched and depleted species, respectively) and computing the average rank for every individual. This non-parametric approach is insensitive to deviations from normal distribution and attributes the highest values to most altered microbiomes. The proportion of cancer cases in different quintiles of the average ranks were computed and the extreme quintiles compared.

---

**Supplementary Fig 1.**
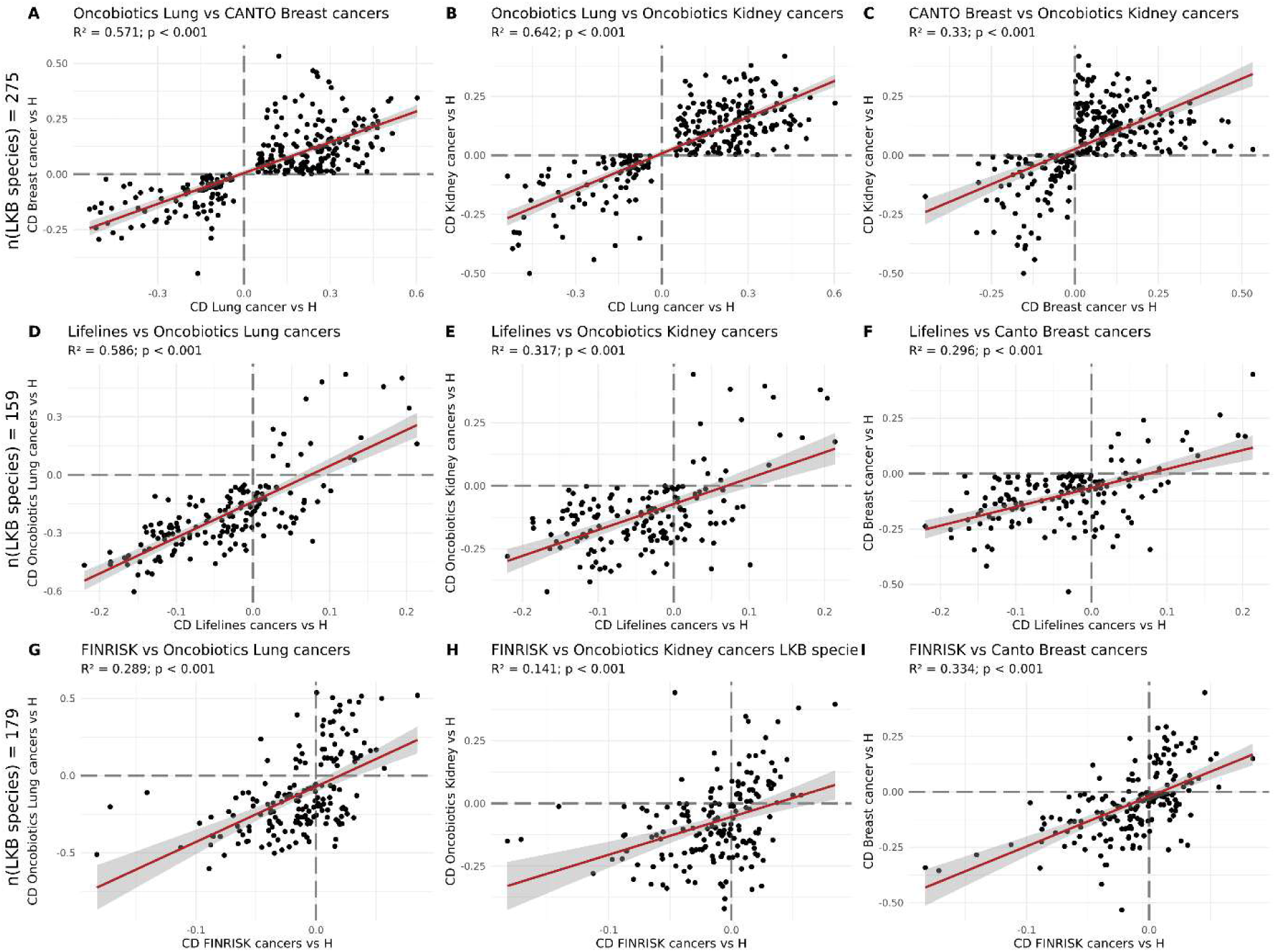
Correlation of Cliff’s deltas for LKB species in different cancer types and cohorts. Cliff deltas were compared pairwise, for all LKB species (n=275) between lung and breast cancers (A), lung and kidney cancers (B) and breast and kidney cancers (C). They were compared pairwise for the LKB species present in the Lifelines cohort (n=159), between all Lifelines prevalent cancers and lung (D), kidney (E) and breast (F) cancers. They were also compared pairwise for the LKB species present in the FINRISK cohort (n=179), between all FINRISK prevalent cancers and lung (G), kidney (H) and breast (I). R^2^ denotes Pearson correlation coefficient squared, p denotes significance of correlation.

**Supplementary Fig 2.**
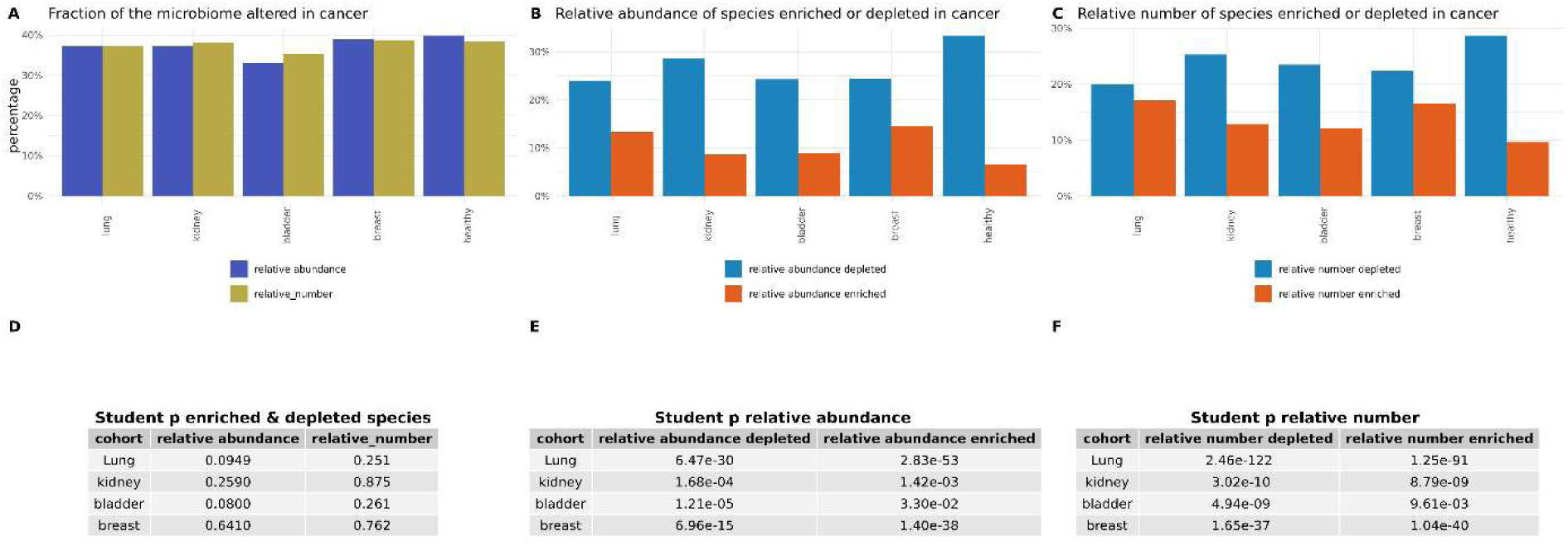
Fraction of the gut microbiome represented by LKB species. (A) Overall relative abundance and number of enriched and depleted species (n=275); (B) Relative abundance of cancer-enriched (n=91) and cancer-depleted (n=184) species; (C) Relative number of cancer-enriched and cancer-depleted (n=184) species. Student’s t-test p-values for overall LKB species fraction (D), relative abundance (E), and relative number (F) in cancer cases compared with healthy individuals.

**Supplementary Fig 3.**
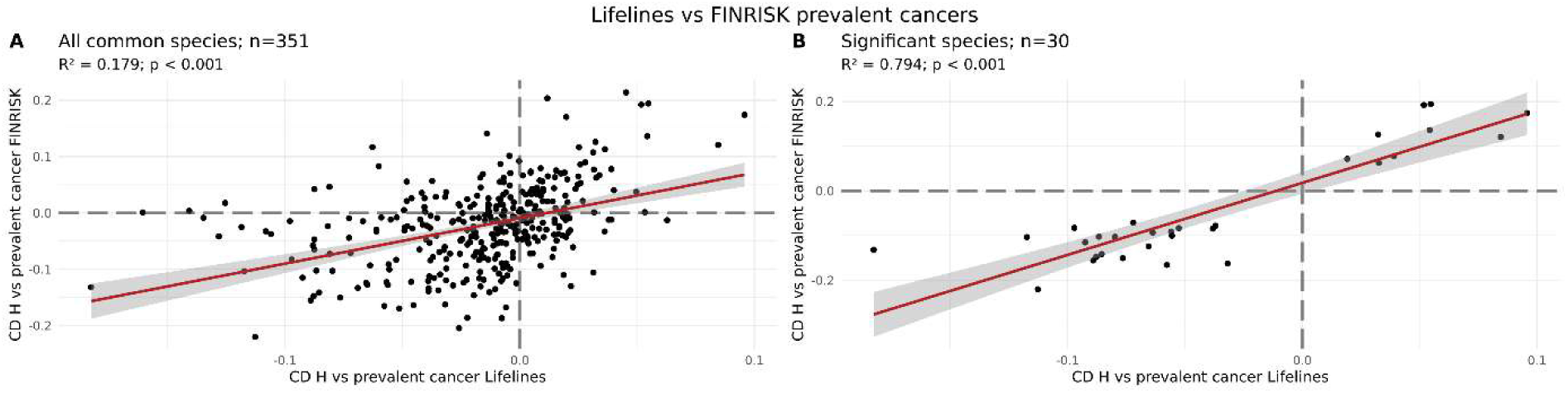
Correlation of Cliff’s deltas for prevalent cancers vs controls in Lifelines and FINRISK cohorts. (A) all common species, (B) species differing significantly in abundance (Wilcoxon p value below 0.05) between cancer cases and controls in both cohorts. R^2^ denotes the squared Pearson correlation coefficient, p denotes significance of correlation, CD denotes Cliff’s delta, H stands for Healthy controls, n indicates number of species used in comparisons.

**Supplementary Fig 4.**
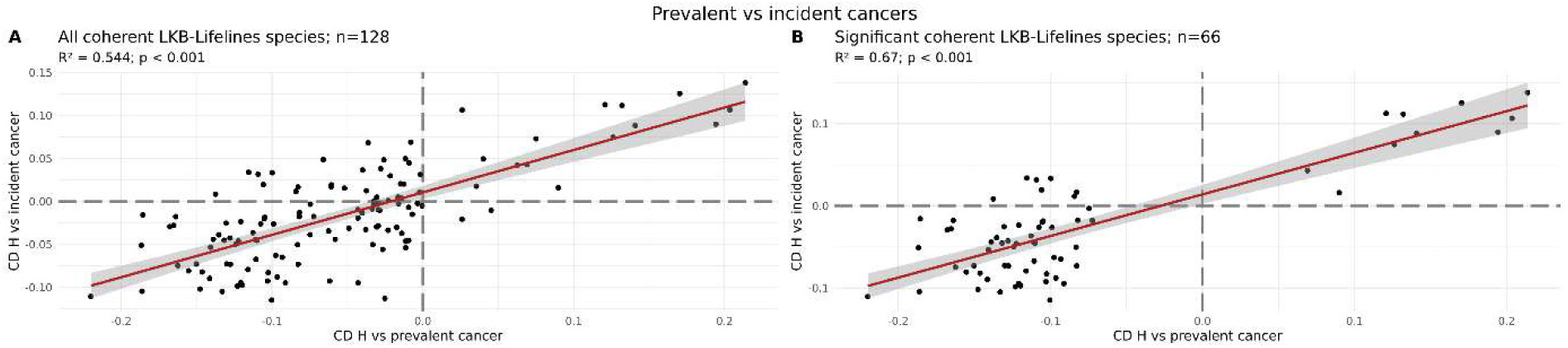
Correlation of Cliff’s deltas for LKB-Lifelines coherent species in prevalent and incident cancers within the Lifelines cohort. (A) Correlation evaluating all LKB species coherent within the Lifelines dataset (n=128), (B) Correlation restricted to the subset of coherent species demonstrating significantly different abundances (Wilcoxon *p* < 0.05) in the healthy-versus-prevalent cancer comparison (n=66). R^2^ denotes the squared Pearson correlation coefficient, p denotes significance of correlation, CD denotes Cliff’s delta, H stands for Healthy controls, n indicates number of species used in comparisons.

**Supplementary Fig 5.**
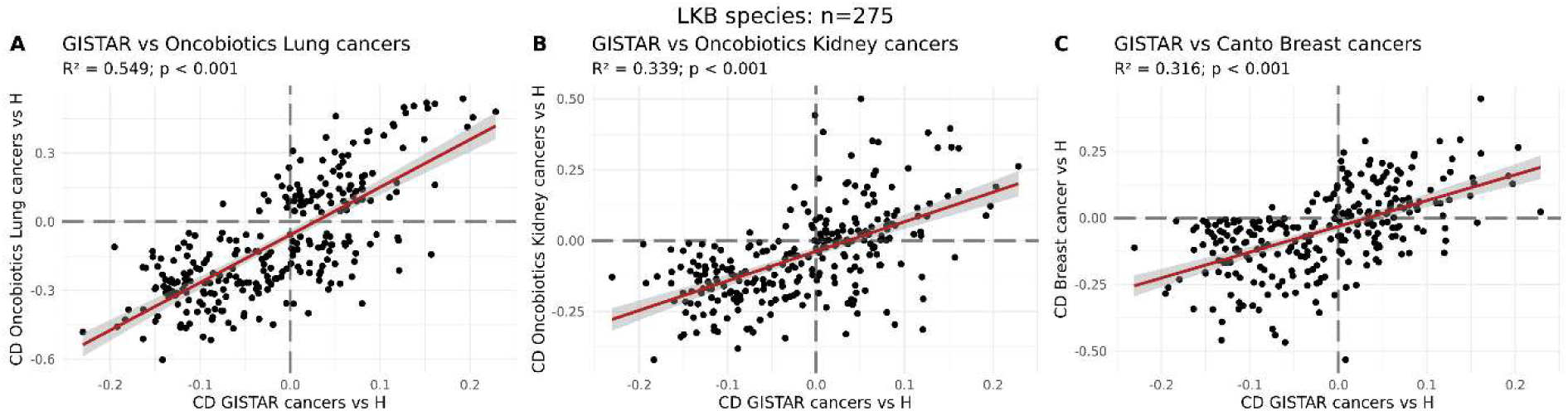
Correlation of Cliff’s deltas between GISTAR incident pan-cancer cases and the three cancer discovery cohorts. Pairwise comparisons were evaluated for all core LKB species (*n* = 275) between GISTAR incident cases versus controls and (A) lung cancer cases, (B) kidney cancer cases, and (C) breast cancers from Oncobiotics and Canto cohorts. R^2^ denotes the squared Pearson correlation coefficient, p denotes significance of correlation, CD denotes Cliff’s delta, H stands for Healthy controls, n indicates number of species used in comparisons.

**Supplementary Fig 6.**
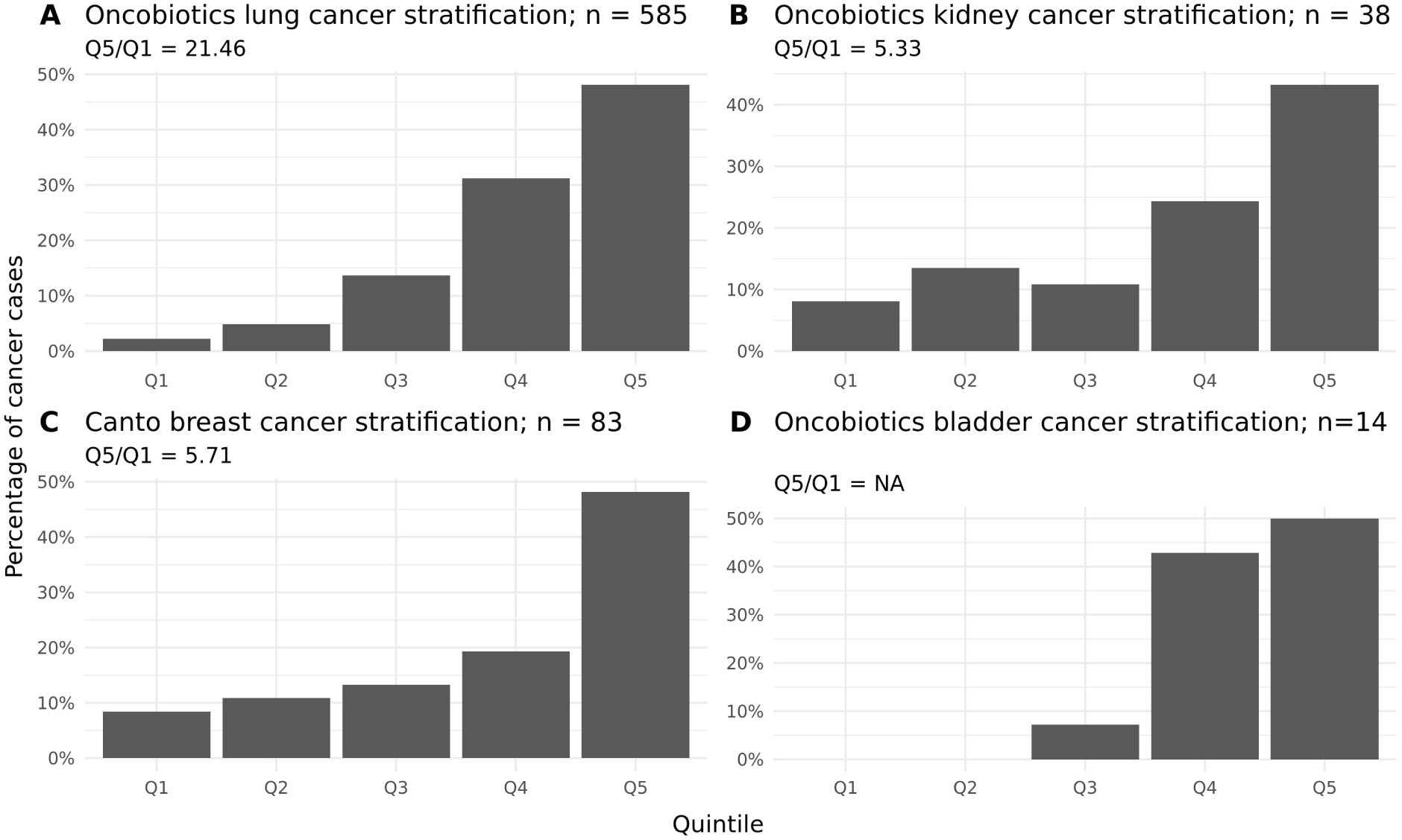
Stratification of distinct clinical cancer cohorts by the level of gut microbiome alterations. Individual microbiome alterations were quantified using the LKB species (*n* = 275). For each comparison, healthy controls from the French Gut cohort (*n* = 5,115) were pooled with individuals from a specific cancer type: lung (*n* = 585; panel A), kidney (*n* = 38; panel B), breast (*n* = 83; panel C), or bladder (*n* = 14; panel D). Each individual was assigned an overall average alteration rank and distributed into the respective quintiles of the rank distribution (Q1 to Q5). The proportion of cancer cases relative to total cancer cases in each quintile is displayed, the risk ratio between the extreme quintiles (Q5/Q1) is shown, n refers to the number of bacterial species used for stratification. For the independent bladder cancer test set (D), the Q5/Q1 ratio was mathematically undefined (NA) due to the complete absence of cancer cases within both the Q1 and Q2 quintiles.

**Supplementary Fig 7.**
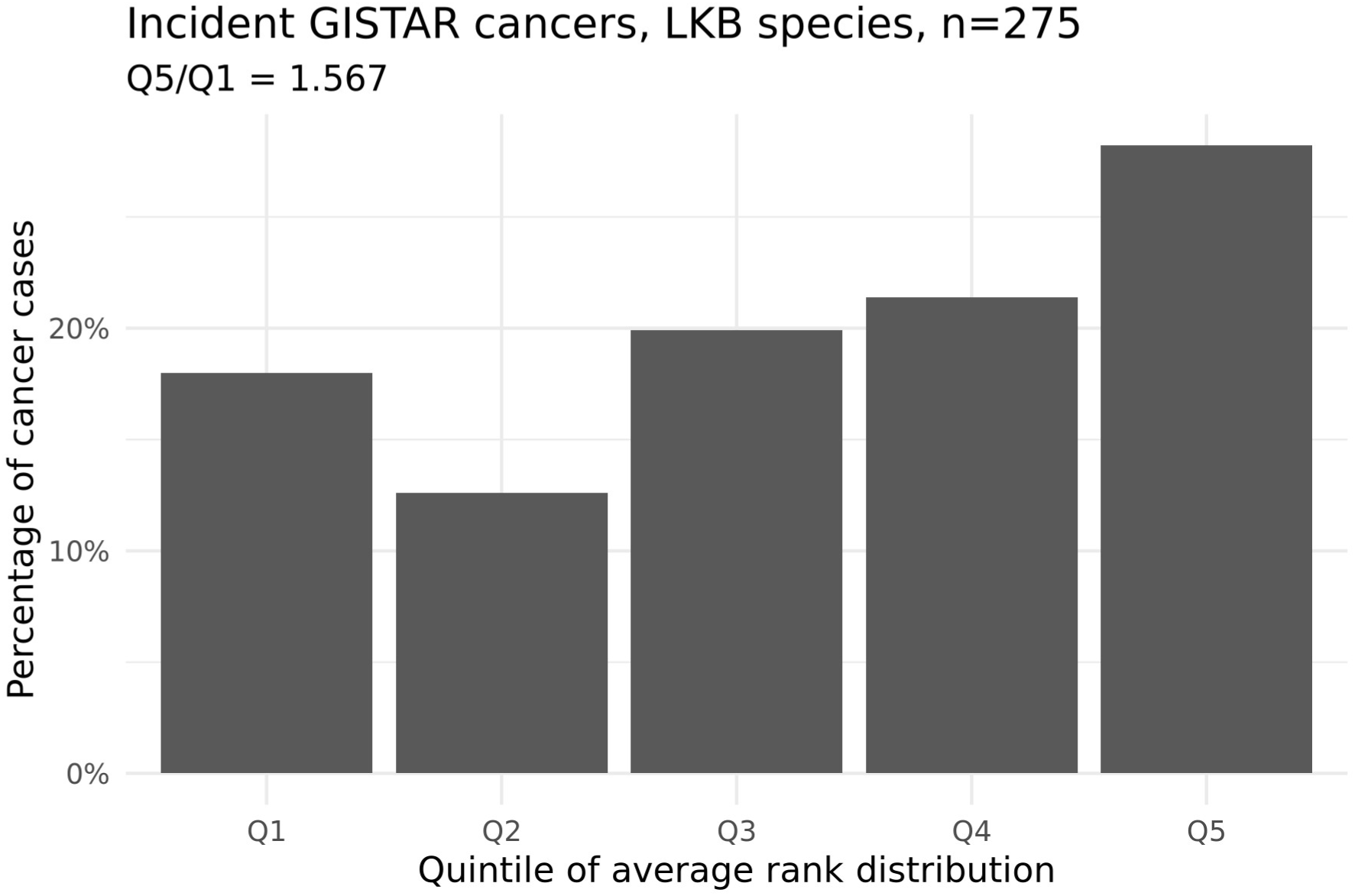
Stratification of the GISTAR incident pan-cancers by the level of gut microbiome alterations. Individual microbiome alterations were quantified using the LKB species (*n* = 275). Each individual within the cohort—encompassing incident cancer cases (*n* = 206) and healthy controls (*n* = 25) was assigned an overall average alteration rank and distributed into the respective quintiles of the rank distribution (Q1 to Q5). The proportion of cancer cases relative to total cancer cases in each quintile is displayed, the risk ratio between the extreme quintiles (Q5/Q1) is shown, n refers to the number of bacterial species used for stratification.

## References

1. Almonte, A. A., Thomas, S., Iebba, V., Kroemer, G., Derosa, L., & Zitvogel, L. (2026). Gut dysbiosis in oncology: A risk factor for immunoresistance. Cell Research, 36(2), 103–120. 10.1038/s41422-025-01212-6

2. Birebent, R., Drubay, D., Alves Costa Silva, C., Marmorino, F., Vitali, G., Piccinno, G., Hurtado, Y., Bonato, A., Belluomini, L., Messaoudene, M., Routy, B., Fidelle, M., Zalcman, G., Mazieres, J., Audigier-Valette, C., Moro-Sibilot, D., Goldwasser, F., Scherpereel, A., Pegliasco, H., … Derosa, L. (2025). Surrogate markers of intestinal dysfunction associated with survival in advanced cancers. Oncoimmunology, 14(1), 2484880. 10.1080/2162402X.2025.2484880

3. Birkeland, E. E., Kværner, A. S., Avershina, E., Bucher-Johannessen, C., Bemanian, V., Blix, H. S., Hjartåker, A., De Vos, W. M., Ursin, G., Hoff, G., Randel, K. R., Botteri, E., Berstad, P., & Rounge, T. B. (2026). Microbiome signatures for detection of colorectal lesions in population-based FIT screening. Nature Communications, 17(1), 8982. 10.1038/s41467-026-75962-1

4. Blanco-Míguez, A., Beghini, F., Cumbo, F., McIver, L. J., Thompson, K. N., Zolfo, M., Manghi, P., Dubois, L., Huang, K. D., Thomas, A. M., Nickols, W. A., Piccinno, G., Piperni, E., Punčochář, M., Valles-Colomer, M., Tett, A., Giordano, F., Davies, R., Wolf, J., … Segata, N. (2023). Extending and improving metagenomic taxonomic profiling with uncharacterized species using MetaPhlAn 4. Nature Biotechnology, 41(11), 1633–1644. 10.1038/s41587-023-01688-w

5. Borodulin, K., Tolonen, H., Jousilahti, P., Jula, A., Juolevi, A., Koskinen, S., Kuulasmaa, K., Laatikainen, T., Männistö, S., Peltonen, M., Perola, M., Puska, P., Salomaa, V., Sundvall, J., Virtanen, S. M., & Vartiainen, E. (2018). Cohort Profile: The National FINRISK Study. International Journal of Epidemiology, 47(3), 696–696i. 10.1093/ije/dyx239

6. Byrd, D. A., Zouiouich, S., Wahl, D., Pardini, B., Gomez Morales, M. F., Tarallo, S., Bulfamante, S., Francavilla, A., Francescato, G., Hogue, S. R., Armaroli, P., Bellisario, C., Ferrante, G., Vogtmann, E., Wan, Y., Hua, X., Shi, J., Gunter, M., Naccarati, A., … Sinha, R. (2026). Fecal immunochemical tests from population-based colorectal cancer screening programs support prospective microbiome cohorts. British Journal of Cancer, 135(6), 931–940. 10.1038/s41416-026-03495-x

7. Cliff, N. (1993). Dominance statistics: Ordinal analyses to answer ordinal questions. Psychological Bulletin, 114(3), 494–509. 10.1037/0033-2909.114.3.494

8. Connan, C., Fromentin, S., Benallaoua, M., Alvarez, A.-S., Pons, N., Quinquis, B., Morabito, C., Nazare, J.-A., Borezée-Durant, E., Le French Gut Consortium, Haimet, F., Ehrlich, S. D., Valeille, K., Cavezza, A., Blottière, H., Veiga, P., Almeida, M., Doré, J., & Benamouzig, R. (2025). Associations Among Diet, Health, Lifestyle, and Gut Microbiota Composition in the General French Population: Protocol for the Le French Gut – Le Microbiote Français Study. JMIR Research Protocols, 14, e64894. 10.2196/64894

9. Coorens, T. H. H., Oh, J. W., Choi, Y. A., Lim, N. S., Zhao, B., Voshall, A., Abyzov, A., Antonacci-Fulton, L., Aparicio, S., Ardlie, K. G., Bell, T. J., Bennett, J. T., Bernstein, B. E., Blanchard, T. G., Boyle, A. P., Buenrostro, J. D., Burns, K. H., Chen, F., Chen, R., … Somatic Mosaicism across Human Tissues Network. (2025). The Somatic Mosaicism across Human Tissues Network. Nature, 643(8070), 47–59. 10.1038/s41586-025-09096-7

10. Derosa, L., Iebba, V., Silva, C. A. C., Piccinno, G., Wu, G., Lordello, L., Routy, B., Zhao, N., Thelemaque, C., Birebent, R., Marmorino, F., Fidelle, M., Messaoudene, M., Thomas, A. M., Zalcman, G., Friard, S., Mazieres, J., Audigier-Valette, C., Sibilot, D. M.-, … Zitvogel, L. (2024). Custom scoring based on ecological topology of gut microbiota associated with cancer immunotherapy outcome. Cell, 187(13), 3373–3389.e16. 10.1016/j.cell.2024.05.029

11. Derosa, L., Routy, B., Fidelle, M., Iebba, V., Alla, L., Pasolli, E., Segata, N., Desnoyer, A., Pietrantonio, F., Ferrere, G., Fahrner, J.-E., Le Chatellier, E., Pons, N., Galleron, N., Roume, H., Duong, C. P. M., Mondragón, L., Iribarren, K., Bonvalet, M., … Zitvogel, L. (2020). Gut Bacteria Composition Drives Primary Resistance to Cancer Immunotherapy in Renal Cell Carcinoma Patients. European Urology, 78(2), 195–206. 10.1016/j.eururo.2020.04.044

12. Derosa, L., Routy, B., Thomas, A. M., Iebba, V., Zalcman, G., Friard, S., Mazieres, J., Audigier-Valette, C., Moro-Sibilot, D., Goldwasser, F., Silva, C. A. C., Terrisse, S., Bonvalet, M., Scherpereel, A., Pegliasco, H., Richard, C., Ghiringhelli, F., Elkrief, A., Desilets, A., … Besse, B. (2022). Intestinal Akkermansia muciniphila predicts clinical response to PD-1 blockade in patients with advanced non-small-cell lung cancer. Nature Medicine, 28(2), 315–324. 10.1038/s41591-021-01655-5

13. Ferlay, J., Ervik, M., Lam, F., Laversanne, M., Colombet, M., Mery, L., Piñeros, M., Znaor, A., Soerjomataram, I., Bray, F. (2024). Global Cancer Observatory: Cancer Today. Lyon, France: International Agency for Research on Cancer. https://gco.iarc.who.int/today/en

14. Fromentin, S., Forslund, S. K., Chechi, K., Aron-Wisnewsky, J., Chakaroun, R., Nielsen, T., Tremaroli, V., Ji, B., Prifti, E., Myridakis, A., Chilloux, J., Andrikopoulos, P., Fan, Y., Olanipekun, M. T., Alves, R., Adiouch, S., Bar, N., Talmor-Barkan, Y., Belda, E., … Pedersen, O. (2022). Microbiome and metabolome features of the cardiometabolic disease spectrum. Nature Medicine, 28(2), 303–314. 10.1038/s41591-022-01688-4

15. Gacesa, R., Kurilshikov, A., Vich Vila, A., Sinha, T., Klaassen, M. A. Y., Bolte, L. A., Andreu-Sánchez, S., Chen, L., Collij, V., Hu, S., Dekens, J. A. M., Lenters, V. C., Björk, J. R., Swarte, J. C., Swertz, M. A., Jansen, B. H., Gelderloos-Arends, J., Jankipersadsing, S., Hofker, M., … Weersma, R. K. (2022). Environmental factors shaping the gut microbiome in a Dutch population. Nature, 604(7907), 732–739. 10.1038/s41586-022-04567-7

16. Ghosh, T. S., Shanahan, F., & O’Toole, P. W. (2022). The gut microbiome as a modulator of healthy ageing. Nature Reviews. Gastroenterology & Hepatology, 19(9), 565–584. 10.1038/s41575-022-00605-x

17. Goel, A., Shete, O., Goswami, S., Samal, A., C B, L., Kedia, S., Ahuja, V., O’Toole, P. W., Shanahan, F., & Ghosh, T. S. (2025). Toward a health-associated core keystone index for the human gut microbiome. Cell Reports, 44(3), 115378. 10.1016/j.celrep.2025.115378

18. Gopalakrishnan, V., Spencer, C. N., Nezi, L., Reuben, A., Andrews, M. C., Karpinets, T. V., Prieto, P. A., Vicente, D., Hoffman, K., Wei, S. C., Cogdill, A. P., Zhao, L., Hudgens, C. W., Hutchinson, D. S., Manzo, T., Petaccia de Macedo, M., Cotechini, T., Kumar, T., Chen, W. S., … Wargo, J. A. (2018). Gut microbiome modulates response to anti-PD-1 immunotherapy in melanoma patients. Science, 359(6371), 97–103. 10.1126/science.aan4236

19. Goulet, L., Plaza Oñate, F., Famechon, A., Quinquis, B., Belda, E., Prifti, E., Le Chatelier, E., & Gautreau, G. (2026). CroCoDeEL: Accurate control-free detection of cross-sample contamination in metagenomic data. Nature Communications, 17(1), 6258. 10.1038/s41467-026-72637-9

20. Grajeda-Iglesias, C., Durand, S., Daillère, R., Iribarren, K., Lemaitre, F., Derosa, L., Aprahamian, F., Bossut, N., Nirmalathasan, N., Madeo, F., Zitvogel, L., & Kroemer, G. (2021). Oral administration of Akkermansia muciniphila elevates systemic antiaging and anticancer metabolites. Aging, 13(5), 6375–6405. 10.18632/aging.202739

21. Gudra, D., Shoaie, S., Fridmanis, D., Klovins, J., Wefer, H., Silamikelis, I., Peculis, R., Kalnina, I., Elbere, I., Radovica-Spalvina, I., Hultcrantz, R., Šķenders, Ģ., Leja, M., & Engstrand, L. (2019). A widely used sampling device in colorectal cancer screening programmes allows for large-scale microbiome studies. Gut, 68(9), 1723–1725. 10.1136/gutjnl-2018-316225

22. Hakozaki, T., Richard, C., Elkrief, A., Hosomi, Y., Benlaïfaoui, M., Mimpen, I., Terrisse, S., Derosa, L., Zitvogel, L., Routy, B., & Okuma, Y. (2020). The Gut Microbiome Associates with Immune Checkpoint Inhibition Outcomes in Patients with Advanced Non-Small Cell Lung Cancer. Cancer Immunology Research, 8(10), 1243–1250. 10.1158/2326-6066.CIR-20-0196

23. Hall, A. B., Yassour, M., Sauk, J., Garner, A., Jiang, X., Arthur, T., Lagoudas, G. K., Vatanen, T., Fornelos, N., Wilson, R., Bertha, M., Cohen, M., Garber, J., Khalili, H., Gevers, D., Ananthakrishnan, A. N., Kugathasan, S., Lander, E. S., Blainey, P., … Huttenhower, C. (2017). A novel Ruminococcus gnavus clade enriched in inflammatory bowel disease patients. Genome Medicine, 9(1), 103. 10.1186/s13073-017-0490-5

24. Jaiswal, S., Fontanillas, P., Flannick, J., Manning, A., Grauman, P. V., Mar, B. G., Lindsley, R. C., Mermel, C. H., Burtt, N., Chavez, A., Higgins, J. M., Moltchanov, V., Kuo, F. C., Kluk, M. J., Henderson, B., Kinnunen, L., Koistinen, H. A., Ladenvall, C., Getz, G., … Ebert, B. L. (2014). Age-related clonal hematopoiesis associated with adverse outcomes. The New England Journal of Medicine, 371(26), 2488–2498. 10.1056/NEJMoa1408617

25. Langmead, B., & Salzberg, S. L. (2012). Fast gapped-read alignment with Bowtie 2. Nature Methods, 9(4), 357–359. 10.1038/nmeth.1923

26. Le Chatelier, E., Almeida, M., Plaza Oñate, F., Pons, N., Gauthier, F., Ghozlane, A., Ehrlich, S. D., Witherden, E., & Gomez-Cabrero, D. (2021). *A catalog of genes and species of the human oral microbiota* [Data set]. Recherche Data Gouv. 10.15454/WQ4UTV

27. Leja, M., Park, J. Y., Murillo, R., Liepniece-Karele, I., Isajevs, S., Kikuste, I., Rudzite, D., Krike, P., Parshutin, S., Polaka, I., Kirsners, A., Santare, D., Folkmanis, V., Daugule, I., Plummer, M., & Herrero, R. (2017). Multicentric randomised study of *Helicobacter pylori* eradication and pepsinogen testing for prevention of gastric cancer mortality: The GISTAR study. BMJ Open, 7(8), e016999. 10.1136/bmjopen-2017-016999

28. López-Otín, C., Blasco, M. A., Partridge, L., Serrano, M., & Kroemer, G. (2023). Hallmarks of aging: An expanding universe. Cell, 186(2), 243–278. 10.1016/j.cell.2022.11.001

29. López-Otín, C., & Kroemer, G. (2021). Hallmarks of Health. Cell, 184(1), 33–63. 10.1016/j.cell.2020.11.034

30. Louca, P., Manning, S., Hackney, E., Sharp, L., Hull, M. A., Koo, S., Young, G. R., Taylor, G. S., Dunneram, Y., Mitra, S., Hampton, J. S., Dobson, C., Neilson, L. J., Addison, C., El-Omar, E. M., the COLO-COHORT research team, Stewart, C. J., & Rees, C. J. (2026). Gut microbiome signatures in colorectal neoplasia: A cross-sectional study across neoplasia stages and subtypes. Gut, gutjnl-2025-337478. 10.1136/gutjnl-2025-337478

31. Meissel, K., & Yao, E. (2024). Using Cliff’s Delta as a Non-Parametric Effect Size Measure: An Accessible Web App and R Tutorial. Practical Assessment, Research, Evaluation Volume 29 Issue 1 2024. 10.7275/PARE.1977

32. Menozzi, E., Ren, Y., Geiger, M., Macnaughtan, J., Avenali, M., Toffoli, M., Gilles, M., Calabrese, R., Mitrotti, P., Gallo, L., Famechon, A., Del Pozo, S. L., Mezabrovschi, R., Koletsi, S., Loefflad, N., Yalkic, S., Limbachiya, N., Clasen, F., Yildirim, S., … Schapira, A. H. V. (2026). Microbiome signature of Parkinson’s disease in healthy and genetically at-risk individuals. Nature Medicine, 32(6), 2096–2106. 10.1038/s41591-026-04318-5

33. Nielsen, H. B., Almeida, M., Juncker, A. S., Rasmussen, S., Li, J., Sunagawa, S., Plichta, D. R., Gautier, L., Pedersen, A. G., Le Chatelier, E., Pelletier, E., Bonde, I., Nielsen, T., Manichanh, C., Arumugam, M., Batto, J.-M., Quintanilha Dos Santos, M. B., Blom, N., Borruel, N., … Ehrlich, S. D. (2014). Identification and assembly of genomes and genetic elements in complex metagenomic samples without using reference genomes. Nature Biotechnology, 32(8), 822–828. 10.1038/nbt.2939

34. Nighot, P., Stine, J., & Clarke, K. (2025). Guts for Self-Eating: Role of Autophagy in Gastrointestinal Health and Disease. Gastro Hep Advances, 4(6), 100654. 10.1016/j.gastha.2025.100654

35. Pennycuick, A., Teixeira, V. H., AbdulJabbar, K., Raza, S. E. A., Lund, T., Akarca, A. U., Rosenthal, R., Kalinke, L., Chandrasekharan, D. P., Pipinikas, C. P., Lee-Six, H., Hynds, R. E., Gowers, K. H. C., Henry, J. Y., Millar, F. R., Hagos, Y. B., Denais, C., Falzon, M., Moore, D. A., … Janes, S. M. (2020). Immune Surveillance in Clinical Regression of Preinvasive Squamous Cell Lung Cancer. Cancer Discovery, 10(10), 1489–1499. 10.1158/2159-8290.CD-19-1366

36. Plaza Oñate, F., & Le Chatelier, E. (2020). *Metagenomic Species Pan-genomes (MSPs) of the human gastrointestinal microbiota* [Data set]. Portail Data INRAE. 10.15454/QVCYRB

37. Plaza Oñate, F., Le Chatelier, E., Almeida, M., Cervino, A. C. L., Gauthier, F., Magoulès, F., Ehrlich, S. D., & Pichaud, M. (2019). MSPminer: Abundance-based reconstitution of microbial pan-genomes from shotgun metagenomic data. Bioinformatics, 35(9), 1544– 1552. 10.1093/bioinformatics/bty830

38. Plaza Onate, F., Pons, N., Gauthier, F., Almeida, M., Ehrlich, S. D., & Le Chatelier, E. (2021). *Updated Metagenomic Species Pan-genomes (MSPs) of the human gastrointestinal microbiota* [Data set]. Recherche Data Gouv. 10.15454/FLANUP

39. Porcari, S., Ng, S. C., Zitvogel, L., Sokol, H., Weersma, R. K., Elinav, E., Gasbarrini, A., Cammarota, G., Tilg, H., & Ianiro, G. (2025). The microbiome for clinicians. Cell, 188(11), 2836–2844. 10.1016/j.cell.2025.04.016

40. Quinquis, B., Famechon, A., Galleron, N., Meslier, V., & Almeida, M. (2024). DNA yield estimation and custom DNA quality evaluation before shotgun metagenomic sequencing. https://www.protocols.io/view/dna-yield-estimation-and-custom-dna-quality-evalua-5qpvo3zzzv4o/v1

41. Routy, B., Gopalakrishnan, V., Daillère, R., Zitvogel, L., Wargo, J. A., & Kroemer, G. (2018). The gut microbiota influences anticancer immunosurveillance and general health. Nature Reviews. Clinical Oncology, 15(6), 382–396. 10.1038/s41571-018-0006-2

42. Routy, B., Le Chatelier, E., Derosa, L., Duong, C. P. M., Alou, M. T., Daillère, R., Fluckiger, A., Messaoudene, M., Rauber, C., Roberti, M. P., Fidelle, M., Flament, C., Poirier-Colame, V., Opolon, P., Klein, C., Iribarren, K., Mondragón, L., Jacquelot, N., Qu, B., … Zitvogel, L. (2018). Gut microbiome influences efficacy of PD-1-based immunotherapy against epithelial tumors. Science, 359(6371), 91–97. 10.1126/science.aan3706

43. Salosensaari, A., Laitinen, V., Havulinna, A. S., Meric, G., Cheng, S., Perola, M., Valsta, L., Alfthan, G., Inouye, M., Watrous, J. D., Long, T., Salido, R. A., Sanders, K., Brennan, C., Humphrey, G. C., Sanders, J. G., Jain, M., Jousilahti, P., Salomaa, V., … Niiranen, T. (2021). Taxonomic signatures of cause-specific mortality risk in human gut microbiome. Nature Communications, 12(1), 2671. 10.1038/s41467-021-22962-y

44. Siegel, R. L., Kratzer, T. B., Wagle, N. S., Sung, H., & Jemal, A. (2026). Cancer statistics, 2026. CA: A Cancer Journal for Clinicians, 76(1), e70043. 10.3322/caac.70043

45. Späth, J., Sewald, Z., Probul, N., Berland, M., Almeida, M., Pons, N., Le Chatelier, E., Ginès, P., Solé, C., Juanola, A., Pauling, J., & Baumbach, J. (2024). Privacy-Preserving Federated Survival Support Vector Machines for Cross-Institutional Time-To-Event Analysis: Algorithm Development and Validation. JMIR AI, 3, e47652. 10.2196/47652

46. Stanley, D., Mason, L. J., Mackin, K. E., Srikhanta, Y. N., Lyras, D., Prakash, M. D., Nurgali, K., Venegas, A., Hill, M. D., Moore, R. J., & Wong, C. H. Y. (2016). Translocation and dissemination of commensal bacteria in post-stroke infection. Nature Medicine, 22(11), 1277–1284. 10.1038/nm.4194

47. Terrisse, S., Derosa, L., Iebba, V., Ghiringhelli, F., Vaz-Luis, I., Kroemer, G., Fidelle, M., Christodoulidis, S., Segata, N., Thomas, A. M., Martin, A.-L., Sirven, A., Everhard, S., Aprahamian, F., Nirmalathasan, N., Aarnoutse, R., Smidt, M., Ziemons, J., Caldas, C., … Zitvogel, L. (2021). Intestinal microbiota influences clinical outcome and side effects of early breast cancer treatment. Cell Death & Differentiation, 28(9), 2778–2796. 10.1038/s41418-021-00784-1

48. Thomas, A. M., Fidelle, M., Routy, B., Kroemer, G., Wargo, J. A., Segata, N., & Zitvogel, L. (2023). Gut OncoMicrobiome Signatures (GOMS) as next-generation biomarkers for cancer immunotherapy. Nature Reviews. Clinical Oncology, 20(9), 583–603. 10.1038/s41571-023-00785-8

49. Yang, X., Kar, S., Antoniou, A. C., & Pharoah, P. D. P. (2023). Polygenic scores in cancer. Nature Reviews Cancer, 23(9), 619–630. 10.1038/s41568-023-00599-x

50. Yonekura, S., Terrisse, S., Alves Costa Silva, C., Lafarge, A., Iebba, V., Ferrere, G., Goubet, A.-G., Fahrner, J.-E., Lahmar, I., Ueda, K., Mansouri, G., Pizzato, E., Ly, P., Mazzenga, M., Thelemaque, C., Fidelle, M., Jaulin, F., Cartry, J., Deloger, M., … Zitvogel, L. (2022). Cancer Induces a Stress Ileopathy Depending on β-Adrenergic Receptors and Promoting Dysbiosis that Contributes to Carcinogenesis. Cancer Discovery, 12(4), 1128–1151. 10.1158/2159-8290.CD-21-0999

51. Yoon, H., Gerdes, L. A., Beigel, F., Sun, Y., Kövilein, J., Wang, J., Kuhlmann, T., Flierl-Hecht, A., Haller, D., Hohlfeld, R., Baranzini, S. E., Wekerle, H., & Peters, A. (2025). Multiple sclerosis and gut microbiota: Lachnospiraceae from the ileum of MS twins trigger MS-like disease in germfree transgenic mice-An unbiased functional study. Proceedings of the National Academy of Sciences of the United States of America, 122(18), e2419689122. 10.1073/pnas.2419689122

52. Zhang, L., & Zhang, J. (2026). Cancer statistics, 2026: Divergent trends and the implementation gap. Nature Reviews Clinical Oncology, 23(4), 237–238. 10.1038/s41571-026-01132-3

53. Zitvogel, L., Derosa, L., Routy, B., Loibl, S., Heinzerling, L., de Vries, I. J. M., Engstrand, L., ONCOBIOME Network, Segata, N., & Kroemer, G. (2025). Impact of the ONCOBIOME network in cancer microbiome research. Nature Medicine, 31(4), 1085–1098. 10.1038/s41591-025-03608-8

54. Zitvogel, L., Fidelle, M., & Kroemer, G. (2024). Long-distance microbial mechanisms impacting cancer immunosurveillance. Immunity, 57(9), 2013–2029. 10.1016/j.immuni.2024.07.020

